# Identification and characterization of genetic risk shared across 24 chronic pain conditions in the UK Biobank

**DOI:** 10.1101/2022.06.28.22277025

**Authors:** Katerina Zorina-Lichtenwalter, Carmen I. Bango, Lukas Van Oudenhove, Marta Čeko, Martin A. Lindquist, Andrew D. Grotzinger, Matthew C. Keller, Naomi P. Friedman, Tor D. Wager

**Affiliations:** Institute of Cognitive Science and Institute for Behavioral Genetics, University of Colorado Boulder, USA; Department of Psychological and Brain Sciences, Dartmouth College, USA; Department of Chronic Diseases and Metabolism, KU Leuven, Belgium; Department of Psychology and Neuroscience and Institute of Cognitive Science, University of Colorado Boulder, USA; Department of Biostatistics, Johns Hopkins University, USA; Department of Psychology and Neuroscience and Institute for Behavioral Genetics, University of Colorado Boulder, USA

**Author notes:** Katerina Zorina-Lichtenwalter, University of Colorado, Institute for Behavioral Genetics, Room 213, 1480 30th St, Boulder, CO 80303, URL: https://www.colorado.edu/ibg, Email addresses.

## Abstract

Chronic pain is attributable to both local and systemic pathology. To investigate the latter, we focused on genetic risk shared among 24 chronic pain conditions in the UK Biobank. We conducted genome-wide association studies (GWAS) on all conditions and estimated genetic correlations among them, using these to model a factor structure in Genomic SEM. This revealed a general factor explaining most of the shared genetic variance in all conditions and an additional musculoskeletal pain-selective factor. Network analyses revealed a large cluster of highly genetically inter-connected conditions, with arthropathic, back, and neck pain showing the highest centrality. Functional annotation (FUMA) showed organogenesis, metabolism, transcription, and DNA repair as associated pathways, with enrichment for associated genes exclusively in brain tissues. Cross-reference with previous GWAS showed genetic overlap with cognition, mood, and brain structure. In sum, our results identify common genetic risks and suggest neurobiological and psychosocial mechanisms of vulnerability to chronic pain.

## 1. Introduction

Chronic pain is a well-documented individual and societal burden, with large costs in suffering [108] as well as cognitive [1], social [26, 46], and economic well-being [97, 77]. These costs are driven by an incomplete understanding of pain chronification mechanisms, which impedes effective prevention and treatment. Chronic pain is often conceptualized as a symptom of a specific, localized pathology. With few exceptions [131], pain conditions are classified based on suspected etiology and/or affected anatomic sites. However, this approach has had limited success. As evidenced by reviews showing that pain treatments fail to reduce pain (e.g. NSAIDs for low back pain [128] and gabapentinoids for neuropathic pain [31]), there is an urgent need for a fundamentally different approach.

A re-conceptualization of chronic pain as a primary disease has been evolving in the pain scholarship community for over 2 decades [93, 111, 101] and recently culminated in the introduction of chronic primary pain disease codes in version 11 of the International Classification of Diseases (ICD-11) [119]. Nevertheless, clinical support for pain sufferers continues to be divided among medical disciplines based on symptoms: back pain is treated by orthopedists, irritable bowel syndrome by gastroenterologists, and so on.

Recent work in the epidemiology of mental health has revealed extensive patterns of co-occurrence across disorders, leading to identification of common factors underlying multiple conditions [88, 53], including the ‘p factor’ [15], which captures general psychopathology. Similar approaches have emerged in pain research, informed by studies of the co-occurrence of pain conditions in large samples [105, 113, 66, 72, 3] and the recognition that different forms of pain are related to similar alterations in the nervous system [65, 76, 54, 63]. Some studies have examined genetic correlations among pain syndromes [129] and genetic risks of having pain in more than 1 of 7 locations on the body [56, 59]. However, a systematic assessment of shared susceptibility across a broad spectrum of pain conditions is lacking, and common factors underlying general pain susceptibility have not yet been characterized.

Most chronic pain conditions are the result of a complex interaction between genetic, environmental, lifestyle, and experiential contributors [80]. However, assessing the genetic component gives insight into commonalities across conditions that can be linked to measurable and potentially targetable biological pathways. In addition, genetic predispositions can be measured cost-effectively using genome-wide association studies (GWASs) and used to characterize and predict risk for chronic pain (e.g., after surgery), predict treatment response, and identify new targets.

We applied this approach here, using the U.K. Biobank (UKBB) [14] and recently developed Genomic Structural Equation Modeling (SEM) software [43]. Genomic SEM enabled us to model the underlying factor structure while accounting for the complex correlations within genetic segments and run a GWAS on the extracted factors. Genomic SEM also produces a Q heterogeneity statistic (QSNP) that indexes single nucleotide polymorphisms (SNPs) unlikely to operate through the genomic factors, such as variants that have a disproportionately strong effect on 1 condition or directionally opposing effects on a subset of traits. Collectively, this allowed for distilling SNPs associated with general pain etiology from trait-specific, genetic pathways. Lastly, we performed functional characterization of common-factor SNPs in FUMA [133].

The questions we aimed to answer in this study were: (1) Is there a general, condition-agnostic genetic risk factor? (2) Are there additional genetic factors underlying subsets of pain conditions? (3) Does the genetic structure correspond to organization of pain by symptom location or hypothesized etiology? (4) What biological pathways and tissues are associated with these genetic factors? Figure 1 provides a graphical overview of the study.

**Figure 1.**
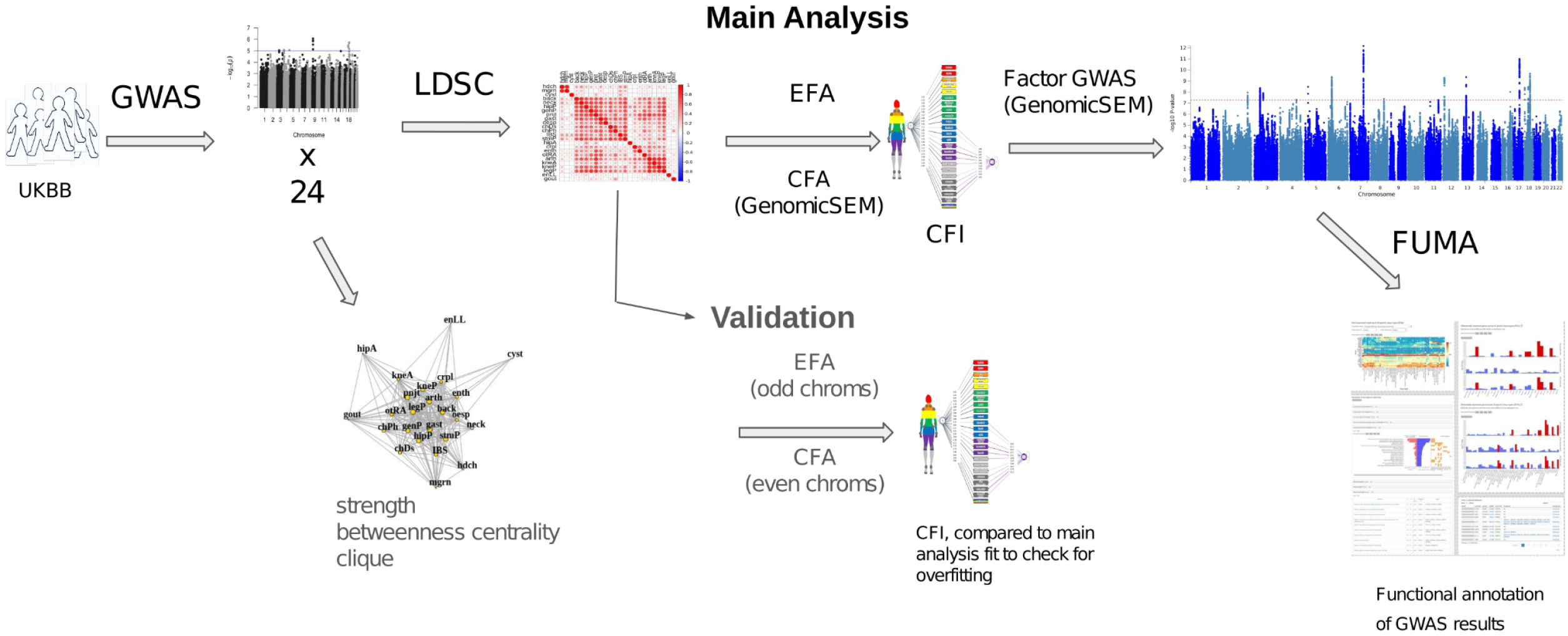
Scheme of study methods and analyses. Abbreviations: GWAS, genome-wide association study; LDSC, linkage-disequilibrium score regression; EFA, exploratory factor analysis; CFA, confirmatory factor analysis; Genomic SEM, genomic structural equation modeling; CFI, comparative fit index.

## 2. Methods

### 2.1 Individual pain conditions

#### 2.1.1 Cohort

Analyses were conducted in the UKBB cohort of participants aged 40-69, who were recruited between 2006 and 2010 (UKBB data-request application 16651). The current standard in genetics is to limit analyses to samples of homogeneous ancestral background to avoid introducing confounds from ethnically mixed samples [115]. We analyzed data from White Europeans (UKBB data field 22006), given that no other group had a sufficient sample size (see Supplementary Table S2 for descriptive statistics of South Asians, the next highest sample size), though analyses in different ancestral groups will be a high priority when more data become available. Individuals who withdrew from the study by August 2020 were removed. A maximum of 435,971 people were included in the analysis, with sample size varying by phenotype, Table 1.

**Table 1:** Pain condition descriptive statistics.

| Con-<br>dition | Full name | Cases | Controls | Preva-<br>lence | $h^2_{SNP}(SE)$ | Reporte<br>d $h^2_{SNP}$ | Citations |
| --- | --- | --- | --- | --- | --- | --- | --- |
| *aCMC | Arthropathy of<br>carpometacarpal joint | 1837 | 201439 | 0.009 | 0.09 (0.036) | NA | NA |
| arth | Arthropathies<br>(non-specific, incl.<br>osteoarthritis) | 80737 | 157458 | 0.339 | 0.09 (0.005) | NA | NA |
| back | Back pain | 119216 | 132641 | 0.473 | 0.09 (0.005) | 0.11/<br>0.12/<br>0.076 | [83,<br>35,<br>114] |
| chDs | Chest pain/discomfort | 72156 | 359415 | 0.167 | 0.08 (0.004) |  |  |
| chPh | Chest pain during<br>physical activity | 2938 | 61044 | 0.046 | 0.13 (0.032) |  |  |
| *Crhn | Crohn's Disease | 1826 | 201030 | 0.009 | 0.13 (0.038) | 0.47 | [121] |
| crpl | Carpal tunnel | 11912 | 424059 | 0.027 | 0.16 (0.011) | 0.02/<br>0.01 | [135,<br>102] |
| CWP | Chronic widespread<br>pain | 6021 | 427884 | 0.014 | 0.14 (0.014) | 0.10 | [56] |
| cyst | Cystitis | 15371 | 189253 | 0.075 | 0.03 (0.008) |  |  |
| *dbNr | Diabetic Neuropathy | 772 | 435199 | 0.002 | 0.13 (0.051) | 0.11 | [85] |
| enLL | Enthesopathies<br>of lower limb | 7000 | 195713 | 0.035 | 0.06 (0.014) |  |  |
| enth | Enthesopathies | 28754 | 175077 | 0.141 | 0.06 (0.007) |  |  |
| *FM | Fibromyalgia | 2149 | 433822 | 0.005 | 0.10 (0.025) | 0.14 | [25] |
| gast | Gastritis | 41746 | 179970 | 0.188 | 0.07 (0.006) |  |  |
| gout | Gout | 15069 | 192253 | 0.073 | 0.20 (0.029) |  |  |
| hdch | Headache | 40222 | 345292 | 0.104 | 0.13 (0.008) | 0.21 | [81] |
| hipA | Hip arthrosis | 17676 | 193048 | 0.084 | 0.14 (0.012) |  |  |
| hipP | Hip pain | 41907 | 381055 | 0.099 | 0.08 (0.005) | 0.12 | [83] |
| IBS | Irritable bowel syndrome | 28419 | 182876 | 0.134 | 0.07 (0.008) |  |  |
| kneA | Knee arthrosis | 31267 | 184763 | 0.145 | 0.14 (0.009) |  |  |
| kneP | Knee pain | 78507 | 334812 | 0.190 | 0.10 (0.005) | 0.08 | [82] |
| legP | Leg pain | 41484 | 108241 | 0.277 | 0.10 (0.008) |  |  |
| mgrn | Migraine | 21586 | 189874 | 0.102 | 0.12 (0.009) | 0.15 | [39] |
| nksh | Neck/Shoulder pain | 72952 | 329192 | 0.181 | 0.08 (0.004) | 0.11 | [84] |
| oesp | Oesophagitis | 13003 | 195329 | 0.062 | 0.06 (0.010) |  |  |
| rhAt | Rheumatoid arthritis | 8685 | 198125 | 0.042 | 0.08 (0.014) |  |  |
| **plr<sub>h</sub> | Polymyalgia rheumatica | 2460 | 433511 | 0.006 | 0.09 (0.023) |  |  |
| pnjt | Pain in joint | 12016 | 423955 | 0.028 | 0.05 (0.008) |  |  |
| *prst | Prostatitis | 3604 | 199950 | 0.018 | 0.06 (0.020) |  |  |
| *seRA | Seropositive rheumatoid arthritis | 839 | 201957 | 0.004 | 0.15 (0.064) |  |  |
| stmP | Stomach pain | 21417 | 396116 | 0.051 | 0.08 (0.006) | 0.14 | [83] |
| **ulcC | Ulcerative colitis | 4211 | 199773 | 0.021 | 0.12 (0.022) |  |  |
| *urCl | Urinary colic | 4743 | 198679 | 0.023 | 0.06 (0.016) |  |  |

$h^2_{\text{SNP}}$ is SNP heritability, variance in the phenotype explained by variance in genotypes (SNPs). S.E. is standard error. Reported $h^2_{\text{SNP}}$ is provided where available. \*phenotypes that did not have a significant $h^2_{\text{SNP}}$ in either the odd or even autosome set.
\*\*phenotypes that did not load significantly onto either the common or specific factor in the EFA-informed CFA.

#### 2.1.2 Phenotypes

The selected phenotypes were either chronic pain conditions, such as migraine or back pain lasting longer than 3 months, or conditions with persistent pain as a prevalent symptom, such as osteoarthritis. An initial list of 92 phenotypes was drawn from 4 UKBB categories: Medical conditions (100074), Health outcomes (713), Self-reported medical conditions (1003), Health and medical history (100036), and First occurrences (1712), downloaded in May 2020. These conditions were recoded into binary phenotypes for analysis (Supplementary Table S1) and subsequently pruned to remove conditions in the following categories: 1. heterogeneous disorders or groupings thereof, such as “Other diabetic polyneuropathies”; 2. branching traits (answers to questions dependent on endorsement of a previous question, with the exception of DF6159: “Pain type(s) experienced in last month”, which was included); 3. disorders with case count *<* 500; 4. disorders that were not sufficiently related to genetics, with SNP heritability less than or equal to 2 standard errors above zero *h*^*2*^_*SNP*_ - 2*SE ≤ 0, as described below. This pruning left 33 conditions (Table 1), further reduced to 24 during the factor analysis step (see section “Factor analysis and Genomic SEM” below).

#### 2.1 3 GWAS

We used genotypes (EGAD00010001474 downloaded using **ukbgene imp**), imputed from the UK10K reference panel [50]. SNP quality control filters consisted of heterozygosity rate (|*F*_het_| >0.2) (determined using the -- het option in Plink), final call rate >0.95 (--geno option in Plink), Hardy-Weinberg equilibrium (1.0×10^−8^), and minor allele frequency >0.01. The sample quality control filter removed mismatches between reported and genotyped sex (Category 100313). We ran GWAS analyses in Regenie [78], using Firth approximation-corrected logistic regression [32]. Briefly, the analysis was run in 2 steps: 1. a model-fitting step, in which genotyped SNPs were split into chunks and 2 levels of ridge regression were run to obtain a per-chromosome genetic predictor of the phenotype; 2. a test of association for all available (imputed) SNPs, also split into chunks, with covariates (described below) regressed out and predictors from the first step removed from phenotypes, using a leave-one-chromosome-out (LOCO) scheme. We used 339,444 genotyped SNPs in 100-SNP chunks in the first step and 11,359,143 imputed SNPs in 200-SNP chunks in the second step, with age, sex, and 10 PCs from genetic PCA (principal component analysis) as covariates [99].

### 2.2 Factor analysis and structural equation model

#### 2.2.1 Heritability and genetic correlations

Using the Genomic SEM R package, summary association statistics from GWAS were converted to z-scores with the **munge** function, and genetic covariances [12] and SNP heritabilities [142] were estimated with the **ldsc** function [13], modified by the authors of Genomic SEM to include a sampling matrix that corrects for sample overlap [43]. We estimated SNP heritability on a continuous liability scale [69] for each pain condition. Genetic covariances were converted to correlations using the **cov2cor** function in R and ordered using hierarchical clustering, which produced a matrix of genetic correlations across pairs of pain conditions (Figure 2A).

**Figure 2.**
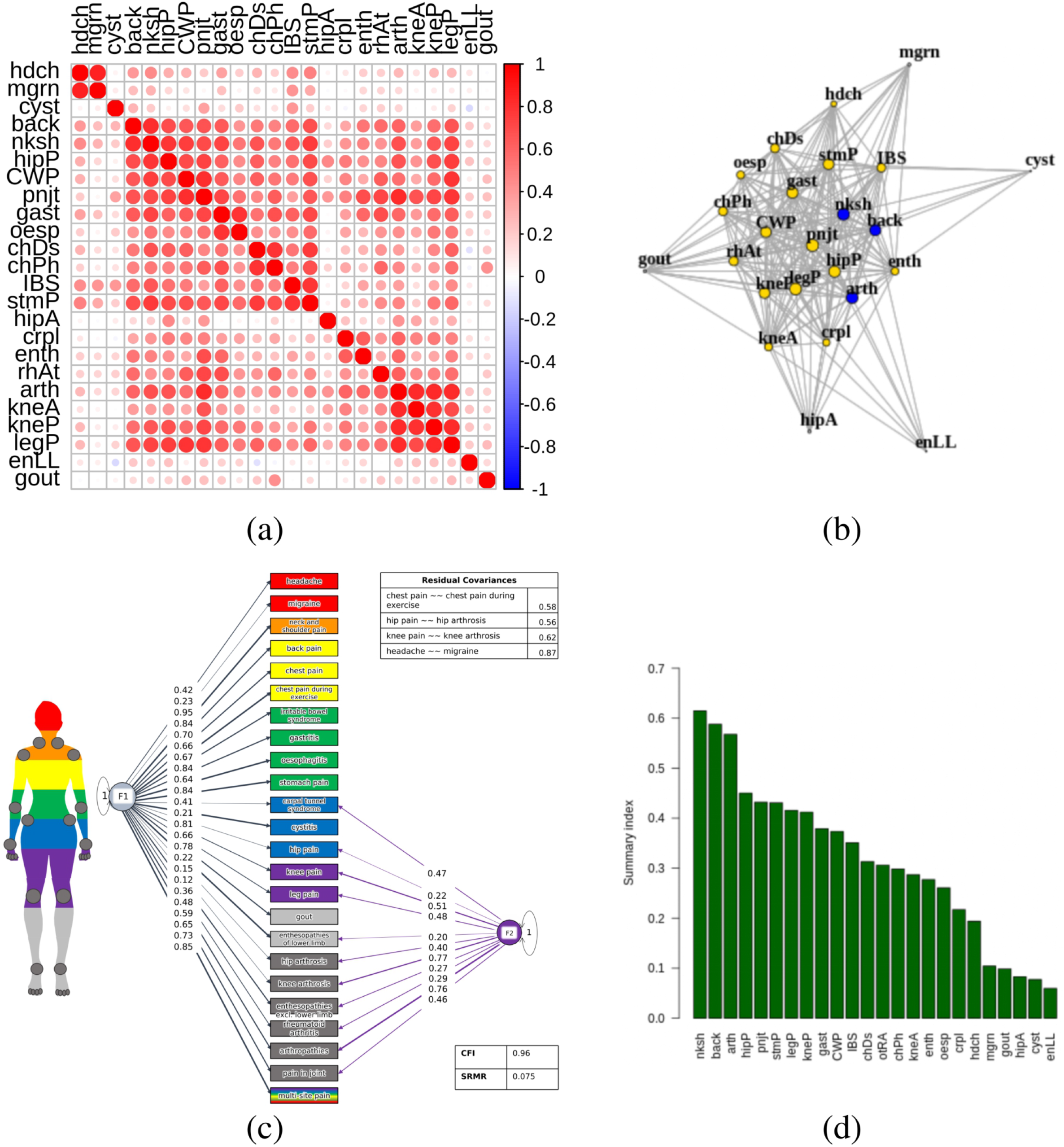
Genetic correlations, network, and Genomic SEM model. (a) Genetic correlations for 24 pain conditions estimated using linkage disequilibrium score regression (LDSC) implemented in Genomic SEM. (b) Network of genetic correlations for 24 pain conditions, pruned for significance at FDR 0.01. The 19 conditions in yellow (arthropathies, back pain, neck/shoulder pain, hip pain, knee pain, leg pain, chest pain (baseline and during physical activity), rheumatoid arthritis, knee arthrosis, joint pain, carpal tunnel, enthesopathies, widespread pain, gastritis, oesophagitis, stomach pain, headache, and IBS) form a clique, complete subgraph. The 3 conditions in blue (neck/shoulder pain, back pain, and arthropathic pain) have the highest betweenness centrality, shortest path between 2 other nodes. Node size corresponds to strength, magnitude-weighted number of connections with other nodes. (c) EFA-CFA model for 24 pain conditions with residual covariances (∼∼) estimated for same body-site conditions: hip arthrosis and pain; knee arthrosis and pain; headache and migraine; chest pain at baseline and during physical activity. F1 is the general factor with positive loadings from all conditions, and F2 is the musculoskeletal factor with positive laodings from carpal tunnel, hip pain, knee pain, leg pain, enthesopathies, rheumatoid arthritis, arthropathies, and pain in joint. CFI, comparative fit index; SRMR, standardized root mean squared residual. All loadings shown are significant at α=0.05. (d) Summary scores (overall measure of interconnectedness for each pain condition) obtained using F1 loadings from EFA-CFA and network strength and betweenness centrality, vector-normalized geometric means (y-axis). (More information on all conditions in Supplementary Table 1 (https://docs.google.com/spreadsheets/d/1S-vFvnwkD5iCP16La_iyjRDTxIqBoMRqAOV6SXpKhN8/edit\#gid=0).

#### 2.2.2 Factor analysis and Genomic SEM

For the factor analytic step, we pursued 3 primary genomic SEM models: one using a confirmatory factor analysis (CFA) informed by an exploratory factor analysis (EFA), and 2 using hypothesis-based CFA. EFA-CFA, is a partially data-driven approach that captures observed groupings in the data, while still permitting structure and inferences based on theory. This approach aligned with our main goal: to test for common factors without rigidly specifying the groupings *a priori*. The 2 hypothesis-based CFAs were used to test whether the observed correlations were well described by pre-specified anatomic and etiological groupings. The anatomic model included the general factor (on which all disorders loaded) and 6 specific factors that group conditions based on body site: Cranial, Gastrointestinal, Joint, Leg/Foot, Pelvic, and Torso. The etiologic model included the general factor and a specific factor for inflammatory conditions, which was the only putative etiology with a substantial number of representative conditions. We discussed a variety of other groupings, but as biological etiology is often unknown – a central problem in pain research – we did not reach clear consensus on additional etiological factors.

For all 3 approaches, the goal was to test a bifactor model, which consisted of a general factor with loadings for all conditions and specific factors that were orthogonal to the general factor and had loadings for specific subsets of conditions. This type of model aligned well with our aim to test whether a common genetic factor underlay all tested conditions, while still allowing for additional shared variance for certain groups of conditions, such as joint-related pain. Similar approaches have been used to model other multidimensional constructs, including personality [18] and psychopathology [9]. EFA as a precursor to CFA has been evaluated in [38] and recently used in [19, 64, 24].

For the EFA portion of the EFA-CFA approach, a scree plot (Supplementary Figure S3) suggested 3 factors. We specified oblique rotation to estimate expected factor covariances. The resulting factor loadings were used to specify a correlated-factors CFA model, first, for testing in Genomic SEM. A condition loaded on a factor in the CFA model if it had positive standardized EFA loadings >0.2, with the highest loading dictating the factor onto which the indicator would load in CFA. If an indicator had 2 loadings within 0.1, both were included in the CFA model. We further specified residual covariances for conditions that were very similar conceptually and had definitional overlap (knee arthrosis and knee pain, hip arthrosis and hip pain, general chest pain/discomfort and chest pain during physical activity, headache and migraine). This obviated one of the 3 factors (a headache-migraine factor), leaving a 2-factor model. Polymyalgia rheumatica and ulcerative colitis were excluded at this step due to lack of positive loadings > 0.2 onto any factor in the EFA model. This correlated factors CFA served as the interim step between EFA and bifactor CFA, in which we specified all conditions to load onto one general factor and the prior model’s correlated factors as uncorrelated additional specific factors.

As the EFA-CFA model is more flexible, we tested for overfitting [33] by repeating the analysis originally conducted on the whole dataset, using a split-genome approach [42, 34]. In step 1, we applied the EFA-CFA procedure described above in odd autosomes (1,3,…21), and in step 2, we assessed the fit of the CFA from step 1 in even autosomes (2,4,…22). This validation step led to a further exclusion of 7 conditions (arthropathy of carpometacarpal joint, diabetic neuropathy, Crohn’s disease, fibromyalgia, prostatitis, seropositive rheumatoid arthritis, and urinary colitis), whose heritability estimates were not significantly above 0 in at least one holdout set, Table 2. This does not imply that these conditions are not heritable or are genetically unrelated to the common factors, as some conditions may be selectively related to genes on odd or even autosomes (limiting replicability here). However, the exclusions helped ensure that the conditions included in the factor model had broad polygenic representation across odd and even autosomes independently. The exclusion left 24 pain conditions for the validation step, which we also used in the main analysis and in the 2 hypothesis-driven approaches for consistency and comparability.

**Table 2:**
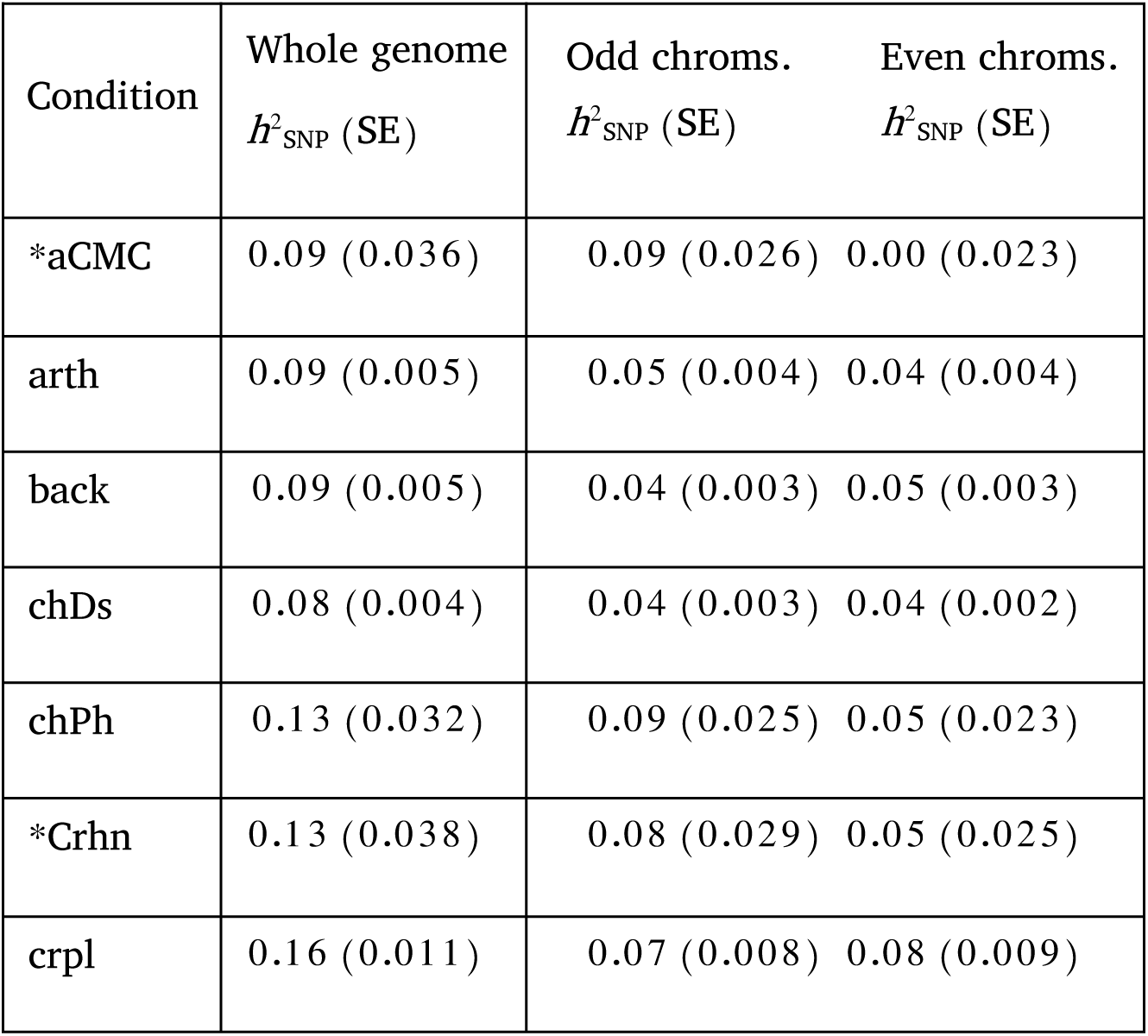
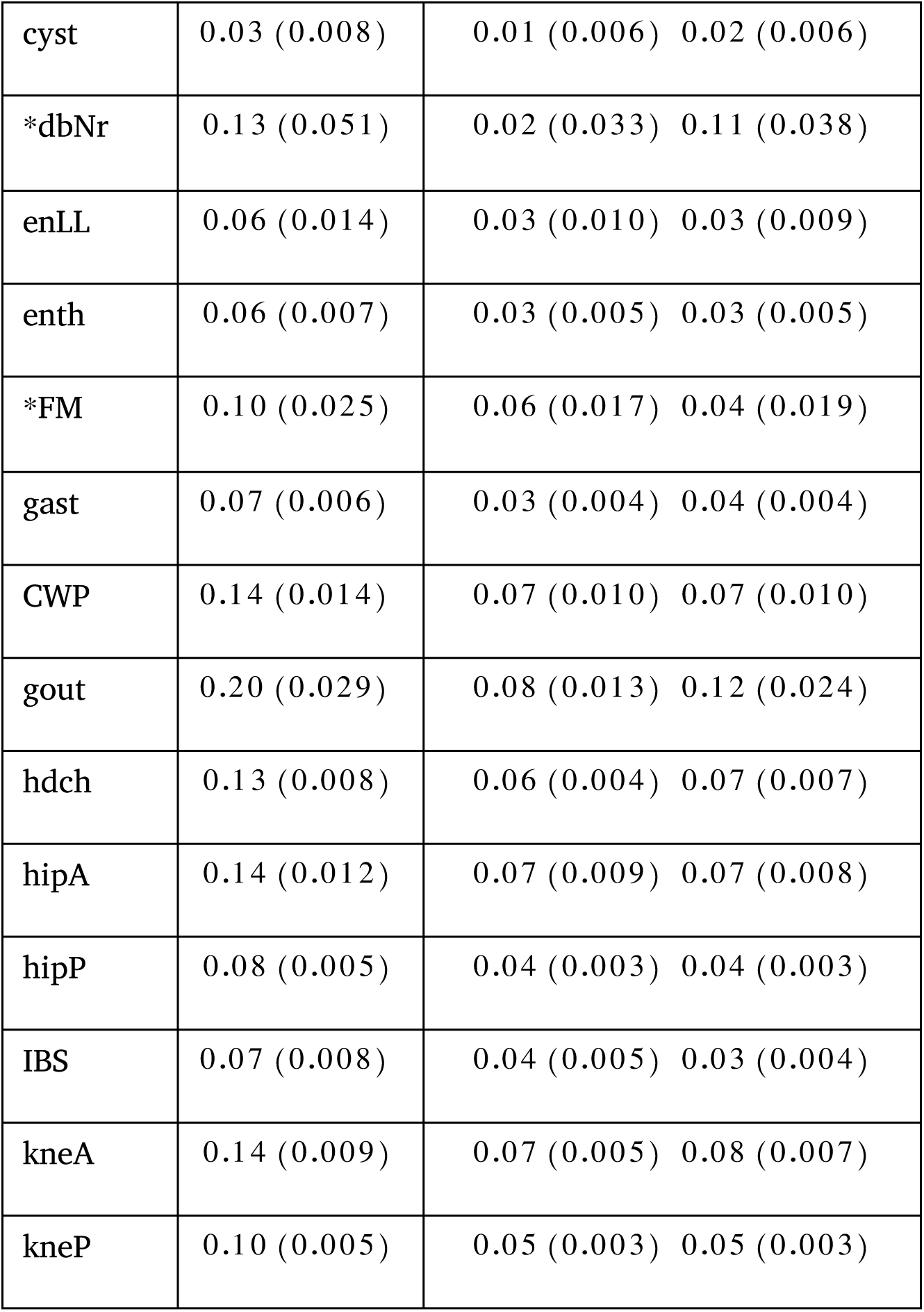
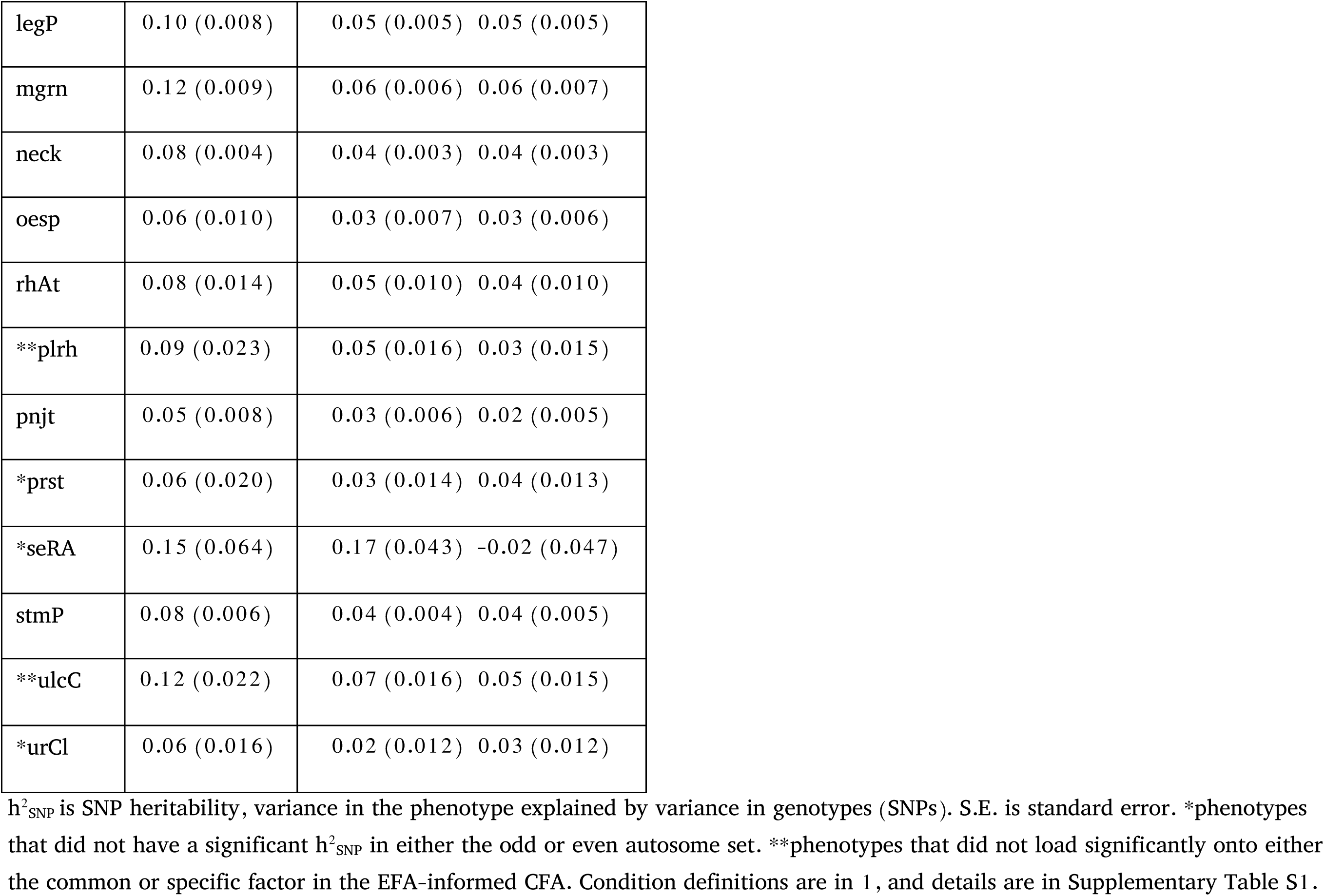
SNP heritability in whole genome, odd and even chromosomes.

| Condition | Whole genome<br>$h^2_{\text{SNP}}$ (SE) | Odd chroms.<br>$h^2_{\text{SNP}}$ (SE) | Even chroms.<br>$h^2_{\text{SNP}}$ (SE) |
| --- | --- | --- | --- |
| *aCMC | 0.09 (0.036) | 0.09 (0.026) | 0.00 (0.023) |
| arth | 0.09 (0.005) | 0.05 (0.004) | 0.04 (0.004) |
| back | 0.09 (0.005) | 0.04 (0.003) | 0.05 (0.003) |
| chDs | 0.08 (0.004) | 0.04 (0.003) | 0.04 (0.002) |
| chPh | 0.13 (0.032) | 0.09 (0.025) | 0.05 (0.023) |
| *Crhn | 0.13 (0.038) | 0.08 (0.029) | 0.05 (0.025) |
| crpl | 0.16 (0.011) | 0.07 (0.008) | 0.08 (0.009) |

|  |  |  |
| --- | --- | --- |
| cyst | 0.03 (0.008) | 0.01 (0.006) 0.02 (0.006) |
| *dbNr | 0.13 (0.051) | 0.02 (0.033) 0.11 (0.038) |
| enLL | 0.06 (0.014) | 0.03 (0.010) 0.03 (0.009) |
| enth | 0.06 (0.007) | 0.03 (0.005) 0.03 (0.005) |
| *FM | 0.10 (0.025) | 0.06 (0.017) 0.04 (0.019) |
| gast | 0.07 (0.006) | 0.03 (0.004) 0.04 (0.004) |
| CWP | 0.14 (0.014) | 0.07 (0.010) 0.07 (0.010) |
| gout | 0.20 (0.029) | 0.08 (0.013) 0.12 (0.024) |
| hdch | 0.13 (0.008) | 0.06 (0.004) 0.07 (0.007) |
| hipA | 0.14 (0.012) | 0.07 (0.009) 0.07 (0.008) |
| hipP | 0.08 (0.005) | 0.04 (0.003) 0.04 (0.003) |
| IBS | 0.07 (0.008) | 0.04 (0.005) 0.03 (0.004) |
| kneA | 0.14 (0.009) | 0.07 (0.005) 0.08 (0.007) |
| kneP | 0.10 (0.005) | 0.05 (0.003) 0.05 (0.003) |
| legP | 0.10 (0.008) | 0.05 (0.005) 0.05 (0.005) |
| mgrn | 0.12 (0.009) | 0.06 (0.006) 0.06 (0.007) |
| neck | 0.08 (0.004) | 0.04 (0.003) 0.04 (0.003) |
| oesp | 0.06 (0.010) | 0.03 (0.007) 0.03 (0.006) |
| rhAt | 0.08 (0.014) | 0.05 (0.010) 0.04 (0.010) |
| **plr<sub>h</sub> | 0.09 (0.023) | 0.05 (0.016) 0.03 (0.015) |
| pnjt | 0.05 (0.008) | 0.03 (0.006) 0.02 (0.005) |
| *prst | 0.06 (0.020) | 0.03 (0.014) 0.04 (0.013) |
| *seRA | 0.15 (0.064) | 0.17 (0.043) -0.02 (0.047) |
| stmP | 0.08 (0.006) | 0.04 (0.004) 0.04 (0.005) |
| **ulcC | 0.12 (0.022) | 0.07 (0.016) 0.05 (0.015) |
| *urCl | 0.06 (0.016) | 0.02 (0.012) 0.03 (0.012) |

Models were evaluated using CFI (comparative fit index), which compares the model fit to one with entirely independent variables, and SRMR (standardized root mean residual), a measure of variance unexplained by the model [49]. A well-fitting model should generally have a CFI≥ .95 and an SRMR≤ .08 [49]. The models were additionally compared using AIC, (Akaike information criterion), a goodness-of-fit index favoring more parsimonious models [60].

#### 2.2.3 Factor GWAS

To estimate the genetic effects of genome-wide variants on the EFA-CFA model factors, we ran a factor GWAS in Genomic SEM (**userGWAS** function). SNP effects on the common factor and one specific factor were calculated by adding the genotypic score for each SNP to the genetic correlation matrix output by **ldsc** for 24 conditions, estimating a new matrix of correlations, and fitting the model with additional paths from the SNP to each of the factors (Supplementary Figure S6).

To conduct a heterogeneity Q test [51, 43], we specified a less restrictive model, in which for every SNP, path coefficients were estimated from the SNP to the individual phenotypes (independent pathways model). We formally assessed the difference between the 2 models – common and independent pathways – using the chi-square difference test. If significant, the SNP’s effects on the individual pain conditions were interpreted to be inadequately modelled by the factor approach. Because this test was calculated for every imputed SNP, we used the standard whole-genome correction for multiple testing, *p <* 5×10^−8^, as the threshold for Q significance. The resulting QSNPs were considered to be associated with pain conditions independent of common factors.

Additionally, we tested for SNPs in linkage disequilibrium (LD), i.e. correlated with QSNPs, using Plink, version 1.9, --indep-pairwise 500 50 0.6, corresponding to 500 kilobases, 50 SNPs, 0.6 *r*^2^ threshold. We removed QSNPs, and SNPs in LD with them, which were liable to capture the contribution of QSNPs, before annotating the GWAS results.

### 2.3 Network analysis

While CFA has many strengths in permitting model comparison, some groups have emphasized that relationships among clinical conditions can have a complex causal structure that can be characterized in terms of networks of interacting variables [127]. We make no strong claims about the underlying causal structure and complement the factor-analytic models with a network-based approach to characterize genetic relationships among conditions. Network characterization and visualization was done in **igraph** in R [22]. Genetic correlations of the final 24 pain conditions were filtered for positive significant correlations, using a threshold of 0.01 false discovery rate (FDR)-corrected, calculated with **fdrtool** in R. We calculated 2 graph-theoretic properties for each pain condition: (1) strength, calculated as the number of edges (genetic correlations with other pain conditions) weighted by their magnitude [7]; and (2) betweenness-centrality, the number of shortest paths between pairs of pain conditions that go through the condition in question) [11]. *Strength* identifies ‘hub’ conditions that are robustly genetically related to many other conditions and may thus be prominent indicators of multi-disorder susceptibility. *Betweenness-centrality* identifies ‘con nector hubs’, conditions that are genetically related to multiple other conditions that are themselves less interrelated. ‘Connector hubs’ are thus key indicators of shared genetic vulnerability. These measures may themselves be correlated, and if so, combined into an overall index, as we did here (described below). At the network level, we estimated the largest *clique*, complete subgraph of intercorrelated pain conditions [29], which identifies a group of genetically linked conditions that may together serve as indicators of multi-disorder susceptibility.

### 2.4 Summary score

To summarize the evidence on which conditions were the most consistent key indicators of multi-disorder vulnerability, we combined results from Genomic SEM and network analysis, obtaining an overall measure of interconnectedness. We derived a summary score for each pain condition using F1 loadings from EFA-CFA, network strength, and betweenness centrality, which are intercorrelated: *r* =.935 for F1 and strength, *r* =.614 for F1 and betweenness, and *r* =.693 for strength and betweenness. We calculated a geometric mean of these 3 measures, after vector-normalizing them using the **norm** function in R.

### 2.5 GWAS annotation

To functionally characterize the genetic contributors to both individual phenotypes and the 2 factors, we submitted all GWAS results to FUMA for prioritization and annotation, using several integrated databases [133]. These analyses consisted of: 1. prioritizing SNPs based on their effect sizes and independence from each other; 2. mapping significant SNPs to genes as described below; 3. conducting a genome-wide gene-based association analysis with FUMA-implemented MAGMA (only used for FUMA’s gene analysis and gene property analyses; 4. gene set analysis for enrichment in known biological pathways; and 5. gene property analysis (testing for preferential expression of associated genes with 53 Gene-Tissue Expression repository (GTEX) tissues). We used standard significance thresholds and parameters, including *p <* 5*e* - 8 for lead SNPs (independent at *r*^2^ < 0.1); *p <* 0.05 for all other SNPs; *r*^2^ threshold for independent significant SNPs used for further annotations, including gene mapping: 0.6; reference panel population = UKB release 2b 10K European; minimum minor allele frequency = 0.01; maximum distance between LD blocks to merge into a locus = 250 kilobases. The *r*^2^ threshold represents a squared pairwise correlation for SNP variant alleles. The sample sizes for the 2 factors (common and musculoskeletal) identified in the final EFA-CFA model were 422,752 and 468,929, respectively, calculated using the method described in [74]. Variants from the reference panel that were in LD with GWAS lead SNPs were included to increase the chance of capturing causal variants.

Mappings of independent significant (as defined in FUMA, *p <* 5×10^−8^ and *r*^2^ *<* 0.6) SNPs onto genes was based on (1) positional distance (within 10 kilobases of gene start and stop coordinates); (2) statistical associations with transcription levels (expression quantitative trait locus, eQTL); and (3) chromatin interaction mapping, physical interactions with gene chromatin states (indicative of transcriptional accessibility). We included protein-coding genes and excluded the major histocompatibility (MHC) region from annotation. MAGMA analysis for gene-based associations [23] was conducted with SNP assignment within windows of 10 kilobases of gene start and stop coordinates, and GTEx, version 8, [71] was used for gene expression analysis in 53 tissues. FUMA parameters are summarized in Supplementary Table S3.

## 3. Results

The work reported here is part of a project preregistered on Open Science Foundation, OSF (Identifying and Characterizing Genetic Susceptibility and Its Overlap with Psychosocial Traits, https://osf.io/4p5e3).

### 3.1 Univariate pain condition GWAS curation and annotation

We considered 24 pain phenotypes in the UK Biobank that (a) were indicative of chronic pain conditions, (b) had sufficient case counts (>500), and (c) were sufficiently heritable (see Methods; Table 1 and Supplementary Table S1). The sample size available for case assessment varied by condition and ranged from 63,982 (chest pain during physical activity) to 435,971 (several conditions). Prevalence ranged from 0.002 (772 cases, diabetic neuropathy) to 0.473 (119,216 cases, back pain). SNP heritability (variance in the phenotype explained by variance in the genotype) ranged from 0.03 (SE 0.008) for cystitis to 0.20 (SE 0.029) for gout.

Summaries of results from univariate GWAS are reported in Table 1 (SNP heritabilities), in Supplementary Figures S1 and S2 for Manhattan and quantile-quantile (QQ)-plots, and in Supplementary Table S14 (numbers of significant SNPs and genes).

### 3.2 Pain condition genetic correlations

Pairwise genetic correlations for the 24 pain conditions, Figure 2A, showed a large cluster of interconnected vertices. This main cluster included etiologically and anatomically diverse conditions, such as back pain, oesophagitis, IBS, and carpal tunnel, suggesting shared genetic susceptibility among these disparate syndromes. Headache and migraine formed a tight mini-cluster (top left), and cystitis, hip arthrosis, enthesopathies of the lower limb and gout showed weaker correlations, suggesting more specific genetic risks for each of these 4 conditions.

### 3.3 Structural equation modeling

Using 3 approaches – hypothesis-driven anatomic (1) and etiologic (2), and largely data-driven exploratory-then-confirmatory (3) factor analyses (EFA-CFA) – we fit a bifactor model to test the loadings of all conditions onto a general factor, with differences in specific factor groupings in each approach. The anatomic model based on body site (Supplementary Figure S4, CFI= .875 and SRMR = .087) and the etiologic model, based on a grouping of inflammatory disorders (Supplementary Figure S5, CFI= .905, SRMR= .095) both had suboptimal fit (CFI≤ 0.95 and SRMR≥ .08), see Methods. The EFA-CFA model, shown in Figure 2C, produced an adequate overall fit (CFI= 0.956, SRMR = 0.075). All pain conditions loaded positively and significantly onto the general factor (F1). The specific factor (F2) had substantial positive loadings for arthropathies (which included osteoarthritis), carpal tunnel, enthesopathies of lower limb, other enthesopathies, hip arthrosis, hip pain, knee arthrosis, knee pain, leg pain, pain in joint, and rheumatoid arthritis.

Given the pronounced musculoskeletal component among these indicators, we interpreted F2 as a musculoskeletal factor. This model was superior (AIC=4849.164) to both the anatomic (AIC=13184.43) and the etiologic (AIC=10024.93) models (see Methods). In addition, the latter models had non-significant loadings on their specific factors (Leg/Foot, Pelvic, and Torso for the anatomic, Supplementary Figure S4, and Inflammatory for the etiologic, Supplementary Figure S5, suggesting that shared variance for those indicators was mainly explained by the general factor (details in Supplementary Note). We validated this model by training on odd (CFI= .884 and SRMR= .123) and testing on even (CFI= 0.903 and SRMR= .129) autosomes (details in Supplementary Note). These comparable metrics in the training, validation, and whole genome datasets suggested that using EFA and CFA on the same dataset did not result in substantial overfitting.

### 3.4 Network analysis and central conditions

Network analysis provided additional evidence for substantial genetic overlap across pain conditions with a different theoretical model (Figure 2B). There was a complete subgraph of 19 interconnected conditions, highlighted in yellow: arthropathies, back pain, neck/shoulder pain, hip pain, knee pain, leg pain, chest pain (baseline and during physical activity), rheumatoid arthritis, knee arthrosis, joint pain, carpal tunnel, enthesopathies, widespread pain, gastritis, oesophagitis, stomach pain, headache, and IBS. Consistent with the CFA model, these conditions affect diverse body sites and span inflammatory and non-inflammatory as well as musculoskeletal and non-musculoskeletal forms of pain. Gout, hip arthrosis, enthesopathies of the lower limb, cystitis, and migraine lay outside the large cluster, but they still had more than 10 connections each. Overall, the network revealed a large core of pain syndromes with shared genetic vulnerability.

Some conditions were particularly central in the network, in several ways. Arthropathies, back, and neck/shoulder pain had the highest betweenness centrality, indicating that genetic associations between many conditions shared genetic vulnerability with at least 1 of these 3.

The summary score derived from F1, network node strength, and betweenness centrality, Figure 2d, reflected the highest degree of genetic overlap with other conditions. Once again, the top highest scorers were neck/shoulder pain, back pain, and arthropathies.

### 3.5 Factor GWAS and annotation

After running factor GWASs, we excluded QSNPs, which showed evidence of effects specific to certain pain conditions (not through the common factors), and we conducted functional annotation of the GWAS output for each of these factors.

#### 3.5.1 General Factor (F1)

The F1 GWAS yielded 33 genome-wide independent significant SNPs, Supplementary Table S4, Figure 3. FUMA mapped these to a total of 241 genes, using at least 1 of 3 methods (positional, eQTL, and chromatin interactions, see Methods), Supplementary Table S5: 26 by positional, 52 by eQTL, and 57 by chromatin interaction mappings. All 3 annotations were identified for 25 genes, highlighted in green in Supplementary Table S5.

**Figure 3.**
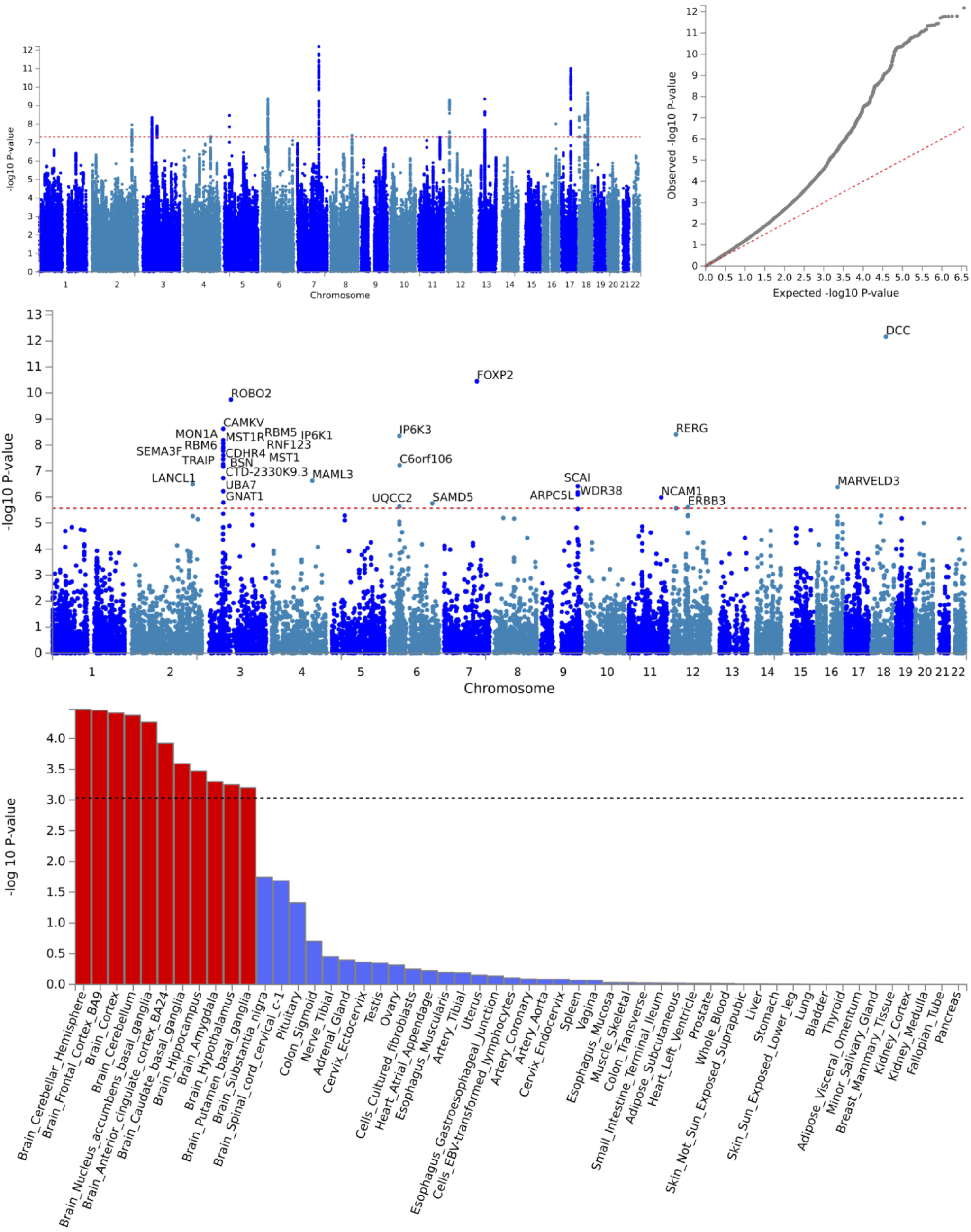
F1 factor GWAS output. Genome-wide association study (GWAS) results for general pain factor (F1). SNP Manhattan (a) and quantile-quantile, QQ, (b) plots for F1 GWAS. (c) Gene-based genome-wide association Manhattan plot, with the top 31 associated genes labelled. (d) Gene property analysis for association between factor GWAS gene effects and gene expression levels in 53 specific tissues from GTEx, version 8.

We used REVIGO [112] to assign, prune, and summarize biological pathways to the 25 genes with overlapping mappings (details in Supplementary Note). The resulting pathways represented by these genes covered a broad range of biological processes, including organ development (gut, heart, muscle and brain), metabolism, catabolism, signaling, immunity, neuronal development, transcription, and DNA repair (Supplementary Table S6).

Additionally, FUMA gene set annotation showed a suggestive significant enrichment (*p* = 1.64×10^−5^, Bonferroni-corrected *p* = .253) for mechanosensory behavior, several neuronal development processes, and several biosynthesis and calcium channel regulation processes, Supplementary Table S7.

MAGMA-based tissue expression analysis, as implemented in FUMA, tested for association between highly expressed genes in 53 GTEx tissues and GWAS effect sizes for the same genes (details in Supplementary Note). Associations were significant only in brain tissues: cortical regions (the cerebral cortex, dorsomedial prefrontal cortex BA9, and anterior cingulate cortex BA24), nucleus accumbens, basal ganglia, amygdala, hippocampus, hypothalamus, and cerebellum, Figure 3D.

Additionally, we used FUMA to cross-reference SNPs and genes with other GWAS reports. Of note was the overlap in SNPs (Supplementary Table S8), and significant enrichment for genes reported to be associated with chronic pain conditions (back pain, Crohn’s disease, IBS, and multi-site chronic pain), brain structural traits, anthropometric traits, cognition and intelligence-related phenotypes, sleep-related phenotypes, neuroticism, and mood phenotypes (Supplementary Figure S9). Genetic overlap with non-pain conditions was suggestive of the complexity of factors contributing to chronic pain. Furthermore, *DCC*, the top gene associated with F1, was also the top gene reported in a recent study of chronic overlapping pain conditions (COPCs), which used pain for more than 3 months in different body sites from the UKBB (head, face, neck/shoulder, back, stomach, hip, knee, all over the body) [59]. Of the 241 genes mapped to independent significant SNPs from the F1 GWAS, *FKBP5* was the only one previously targeted in a candidate gene study (as opposed to GWAS) for posttraumatic musculoskeletal pain [144, 10].

#### 3.5.2 Musculoskeletal Factor (F2)

The F2 GWAS yielded 7 genome-wide significant lead SNPs, Supplementary Table S9. Positional mapping yielded 5 unique genes; eQTL mapping yielded 18 genes; and chromatin interaction mapping yielded 19 genes, with 5 genes mapped using all 3 methods, green: *DPYD, MAPK6, GLIS3, COL27A1, and SLC44A2*, Supplementary Table S10.

REVIGO pathway analysis showed associations with genes involved in bone and neuronal development, cell cycle, transcription regulation and signal transduction, Supplementary Table S11. Gene set annotation showed a Bonferroni-corrected significant enrichment for regulation of RNA biosynthetic process and nominally significant (*p <* 0.05) enrichment for several other regulatory processes, chromatin organization, cell migration involved in heart development, and DNA damage response, Supplementary Table S12. MAGMA tissue expression analysis found no significant association between gene expression and GWAS effect sizes for 53 tissues, Supplementary Figure S7D.

Cross-referencing with other GWAS reports identified previously reported SNP associations with anthropometric traits (height, hip circumference, offspring birth weight), hip or knee osteoarthritis, sleep-related phenotypes, and type 2 diabetes (Supplementary Table S13), and significant overlap with genes reported to be associated with inflammatory skin disease, palmitic and stearic acid levels, (Supplementary Figure S9). None of the genes previously targeted in candidate gene studies for pain [144] mapped to independent significant SNPs for F2.

## 4. Discussion

The UKBB is a large and extensively phenotyped cohort, which recently added First Occurrences data (category 1712), giving researchers access to primary care and death register records to supplement self-reports and ICD-10 diagnoses, earlier available exclusively from hospital intake records. This growing trove of genotypic and phenotypic data has enabled us to examine a much larger number of pain conditions than reported in prior genetic studies of multi-site pain [58, 56, 59, 120]. Our work builds on earlier studies, which included smaller numbers of conditions, often selected *a priori* based on anatomic proximity or hypothesized etiology. Most of these have been conducted in twins. They include reports of genetic correlations of musculoskeletal pain in different body sites [136]; spinal pain syndromes [45]; chronic pain syndromes (chronic widespread musculoskeletal pain, chronic pelvic pain, migraine, and IBS), which estimated 66% heritability using a common pathway model [130]; low back pain with common widespread pain [73]; and TMD with migraine [98].

Several large-scale chronic pain GWAS, with strengths complementary to twin studies [36], have been published on pain in the past 3 years [114, 81, 35, 82, 84, 86]. These reports, which used earlier releases of the UKBB, include 3 on multi-site pain [56, 57, 59]. However, a significant caveat to earlier GWAS on multi-site pain as a single phenotype is the potential for gene variants that selectively act on one of the conditions included in the multi-site definition to be interpreted as cross-condition variants. The Genomic SEM approach has allowed us to extract genetic variance that is truly common to different conditions, rather than detecting associations with an averaged phenotype. In other words, it enabled us to capture genetic risk for chronic pain, regardless of etiology or symptomatology.

Our results identified a single common genetic factor that explained substantial genotypic variance in pain conditions with different suspected etiologies and anatomic presentations, as evidenced by significant loadings onto this factor for all 24 conditions tested. This common factor implied shared genetic risk for a range of conditions, some as clinically distinct as migraine and cystitis, and pointed to their shared systemic pathophysiology. Additionally, a second factor explained some of the shared genetic variance across diverse musculoskeletal conditions: arthropathies, carpal tunnel, enthesopathies (general and lower limb), hip arthrosis, hip pain, knee arthrosis, knee pain, leg pain, and joint pain. This musculoskeletal factor was in line with the World Health Organization’s grouping of pain diseases of the musculoskeletal system, which groups conditions that affect joints, bones, muscles, the spine, and multiple body areas or systems [134]. The 2-factor model explained the pattern of genetic associations among disorders better than either the anatomic or etiologic grouping of known inflammatory disorders. The shared genetic burden was also apparent in network-based analyses, which complemented factor analyses by conceptualizing common risk in terms of multiple local causes instead of a few latent causes.

The existence of widespread shared genetic risk factors – and the existence of a general factor in particular – challenges the current clinical practice of grouping chronic pain conditions based on location of symptoms on the body or suspected etiology [30]. Evidence for central processes beyond local pathophysiology has been accumulating, including recent studies demonstrating that chronic pain is best conceptualized as a combination of biopsychosocial factors that may lead to a variety of pain conditions [21, 125, 75, 20, 124, 16]. Studies in rodents have identified neuroinflammation- and neuroplasticity-related changes in brain pathways that mediate persistent pain behavior in animal models of different pain modalities [143, 96, 91, 27, 6, 122, 52, 5, 37, 87]. Neuroimaging studies have identified common brain systems involved in musculoskeletal pain [79, 68, 17, 44, 67, 61, 90], IBS [106, 4], orofacial pain [2], neuropathic pain [95, 100, 139], and postsurgical pain [47]. These different lines of evidence have led to a new classification system for chronic pain in ICD-11.

The new system shifts pain category assignment from “perceived location”, “etiology”, or “primarily affected anatomical system” to a hierarchical approach, which assigns based on etiology first, then pathophysiology, then body site, and allows for “multiple parenting”, i.e. assignment of a diagnosis to multiple categories [118]. Furthermore, chronic primary pain was included in the ICD-11 as a diagnosis “agnostic with regard to etiology” [92]. These changes are important steps toward a more comprehensive characterization of chronic pain that considers the complex and multifaceted nature of its experience.

In service of this goal, our model suggests that, in addition to condition-specific genetic susceptibility, there is a genetically encoded pathophysiology common to different chronic pain conditions. This supports the view of chronic pain as a disorder involving systemic pathology and of localized peripheral pain as a possible symptom of non-local vulnerabilities and central pathophysiology [137].

Beyond identifying shared genetic risk components, the functional annotation of these components offers insight into possible molecular mechanisms involved in their pathophysiology. Our study adds to existing evidence for the role of *DCC*, which is involved in axonal guidance [140], in chronic pain [114, 103, 59]. However, the presence of other statistically significantly associated genes and the myriad different pathways they tag suggests that focus exclusively on the top association – however tempting it may be to uphold this representative of the nervous system in a pain study – may be detrimental to obtaining a more comprehensive picture of pain susceptibility. Primary chronic pain, like many other complex traits, is very likely a highly polygenic trait [138]. Nor is the highest association statistic necessarily correlated with its relative importance (see this recent publication discussing negative selection as a mechanism for purging high-effect variants in critical genetic loci [94]). Therefore, our approach is to prioritize genes based on: 1. an *a priori* association *p* value cut-off to ensure statistical rigor; and 2. convergent lines of evidence for functional importance, i.e. overlap in mapping approaches. In the resulting set, we interpret our findings in their entirely, without deference to the top association.

Gene mapping of the common factor (F1) implicates a large number of genes. Gene set analysis highlights genes with regulatory function and likely pleiotropy, i.e., roles in other complex traits [132]. There is converging evidence for the involvement of the nervous system: gene expression data shows an enrichment exclusively for brain tissues, and FUMA gene set analysis implicates biological processes specific to the nervous system. Echoing a recent report of heritability enrichment for chronic overlapping pain conditions exclusive to the CNS [59], these findings provide a genetic line of evidence for the reported alterations in brain circuitry shared by chronic pain conditions [116, 5, 62, 52]. In addition to CNS activity, however, the pathways mapped in FUMA implicate a broad range of other functions, such as gut development, locomotion, and protein secretion, suggesting that susceptibility to chronic pain may involve other systemic biological changes. The overlap with genetic variants previously reported in GWAS for cognitive, structural, mood, and personality traits, regulation of inflammation and neuroplasticity, and psychiatric disorders underscores the highly multifaceted nature of pain as a biopsychosocial condition, while providing new clues about the key genes and systems involved [21, 125, 75, 20, 124, 16].

As might be expected, the genes associated with the specific musculoskeletal factor are fewer, and their pathways are less diverse. They implicate skeletal development, choline transport, signaling, and transcription machinery. Notably, they do not implicate the nervous system. Overlap with previous GWAS results suggests involvement of variants affecting anthropometric traits, thus implicating body-structural mechanisms. Similar associations have been shown for musculoskeletal pain conditions before: genetic overlap in osteoarthritis with height and BMI [28], back pain [35] and multi-site musculoskeletal pain [120] with structural-anatomic genes.

### 4.1 Limitations

There are several notable limitations of this study. First, the precise biological and psychological traits that may underlie the common genetic pain factors remain to be elucidated in future studies. The annotated genetic profile of the general factor (F1) suggests a combination of systemic biological and psychological predispositions, including the tendency to evaluate somatic experience in a more negative way (genetic association overlap with traits such as neuroticism and moodiness). Prior studies of psychosocial [48], biological [107], as well as structural and functional brain [126, 63] correlates for pain should be extended to assess the specific roles of each of these contributors to general pain risk, as was recently done for chronic back pain [123].

The second limitation lies in the reliance of annotations obtained from FUMA on the information available in existing data repositories, which may be restricted by insufficient resolution or small sample sizes. Thus, although we did not find associations with inflammatory cytokines, a wealth of evidence links inflammation with multiple forms of chronic pain [110, 55, 40] and should be investigated further.

Third, the genetic scores of our common factors should be validated for association with chronic pain or intermediate phenotypes using either association analysis in an independent sample or cross-validation methods. This would require obtaining a polygenic score and is the aim of a follow-up study.

Fourth, given sample size limitations in the UKBB for non-European individuals, we were not able to test our model for generalizability across ancestral populations. On a related note, it would be of interest to examine the common factors for sex differences, given a larger sample.

By establishing genetic risk factors in a large sample, this study paves the way for more detailed assessments of pain prognosis and treatment response in targeted studies. For example, the ongoing Acute to Chronic Pain Signatures (A2CPS) study aims to establish risk factors for post-surgical pain from genetic, multi-omics, psychosocial, and neuroimaging measures in another large sample (3,000 patients; a2cps.org). Our factor scores could be tested as prognostic risk factors for chronic post-surgical pain, combined with psychosocial [104, 89] and quantitative sensory testing (QST) measures [30, 119, 109].

### 4.2 Conclusions

In summary, our findings support the hypothesis that there is a genetic susceptibility common to a broad range of diverse chronic pain conditions. The shared pathophysiology for the conditions examined here appears to lie partly in the CNS and partly scattered across many different systems and functional processes. Additionally, there is a body-wide, suggestively musculoskeletal system-specific shared genetic factor. These findings are consistent with emerging views that chronic pain is a disease in its own right [116, 92, 124], meaning that one systemic pathology underlies disparate types of pain. Our results help identify and characterize the genetic components of this pathology and suggest that brain prefrontal and affective/motivational circuits may play a key role, supporting converging evidence from animal [117, 54, 141, 41, 52] and human [8, 91, 62, 70] studies. Together, this evidence underscores the importance of new ways to diagnose and treat chronic pain, whereby a given chronic pain condition is not considered as only a symptom of a localized somatic disease but is seen as a manifestation of an underlying shared pathology with concurrent risk for other pain conditions and previously unexplored centralized treatment targets.

## Data Availability

All data produced in the present study are available upon reasonable request to the authors.

## Acknowledgements

This work is supported by R01DA046064, “Brain and Genetic Predictors of Individual Differences in Pain and Placebo Analgesia”; PIs are Naomi Friedman and Tor Wager. The authors declare no conflict of interest.

## References

1. Apkarian, A. V., Sosa, Y., Krauss, B. R., Thomas, P. S., Fredrickson, B. E., Levy, R. E., Harden, R. N. and Chialvo, D. R. [2004], ‘Chronic pain patients are impaired on an emotional decision-making task’, Pain 108(1-2), 129–136.

2. Ayoub, L. J., Seminowicz, D. A. and Moayedi, M. [2018], ‘A meta-analytic study of experimental and chronic orofacial pain excluding headache disorders’, NeuroImage: Clinical 20, 901–912.

3. Bair, E., Gaynor, S., Slade, G. D., Ohrbach, R., Fillingim, R. B., Greenspan, J. D., Dubner, R., Smith, S. B., Diatchenko, L. and Maixner, W. [2016], ‘Identification of clusters of individuals relevant to temporomandibular disorders and other chronic pain conditions: the OPPERA study’, Pain 157(6), 1266.

4. Baliki, M. N., Baria, A. T. and Apkarian, A. V. [2011], ‘The cortical rhythms of chronic back pain’, Journal of Neuroscience 31(39), 13981–13990.

5. Baliki, M. N., Mansour, A. R., Baria, A. T. and Apkarian, A. V. [2014], ‘Functional reorganization of the default mode network across chronic pain conditions’, PloS One 9(9), e106133.

6. Bannister, K. and Dickenson, A. [2017], ‘The plasticity of descending controls in pain: translational probing’, The Journal of physiology 595(13), 4159–4166.

7. Barrat, A., Barthelemy, M., Pastor-Satorras, R. and Vespignani, A. [2004], ‘The architecture of complex weighted networks’, Proceedings of the national academy of sciences 101(11), 3747–3752.

8. Becker, S., Navratilova, E., Nees, F. and Van Damme, S. [2018], ‘Emotional and motivational pain processing: Current state of knowledge and perspectives in translational research’, Pain Research & Management.

9. Bornovalova, M. A., Choate, A. M., Fatimah, H., Petersen, K. J. and Wiernik, B. M. [2020], ‘Appropriate use of bifactor analysis in psychopathology research: Appreciating benefits and limitations’, Biological Psychiatry 88(1), 18–27.

10. Bortsov, A. V., Smith, J. E., Diatchenko, L., Soward, A. C., Ulirsch, J. C., Rossi, C., Swor, R. A., Hauda, W. E., Peak, D. A., Jones, J. S. et al. [2013], ‘Polymorphisms in the glucocorticoid receptor co-chaperone FKPB5 predict persistent musculoskeletal pain after traumatic stress exposure’, Pain 154(8), 1419–1426.

11. Brandes, U. [2001], ‘A faster algorithm for betweenness centrality’, Journal of mathematical sociology 25(2), 163–177.

12. Bulik-Sullivan, B., Finucane, H. K., Anttila, V., Gusev, A., Day, F. R., Loh, P.-R., Duncan, L., Perry, J. R., Patterson, N., Robinson, E. B. et al. [2015], ‘An atlas of genetic correlations across human diseases and traits’, Nature genetics 47(11), 1236.

13. Bulik-Sullivan, B. K., Loh, P.-R., Finucane, H. K., Ripke, S., Yang, J., Patterson, N., Daly, M. J., Price, A. L. and Neale, B. M. [2015], ‘LD score regression distinguishes confounding from polygenicity in genome-wide association studies’, Nature genetics 47(3), 291–295.

14. Bycroft, C., Freeman, C., Petkova, D., Band, G., Elliott, L. T., Sharp, K., Motyer, A., Vukcevic, D., Delaneau, O., OConnell, J. et al. [2018], ‘The UK Biobank resource with deep phenotyping and genomic data’, Nature 562(7726), 203–209.

15. Caspi, A., Houts, R. M., Belsky, D. W., Goldman-Mellor, S. J., Harrington, H., Israel, S., Meier, M. H., Ramrakha, S., Shalev, I., Poulton, R. et al. [2014], ‘The p factor: one general psychopathology factor in the structure of psychiatric disorders?’, Clinical psychological science 2(2), 119–137.

16. Cay, M., Gonzalez-Heydrich, J., Teicher, M. H., van der Heijden, H., Ongur, D., Shinn, A. K. and Upad-hyay, J. [2022], ‘Childhood maltreatment and its role in the development of pain and psychopathology’, The Lancet Child & Adolescent Health.

17. Ceko, M., Shir, Y., Ouellet, J. A., Ware, M. A., Stone, L. S. and Seminowicz, D. A. [2015], ‘Partial recovery of abnormal insula and dorsolateral prefrontal connectivity to cognitive networks in chronic low back pain after treatment’, Human brain mapping 36(6), 2075–2092.

18. Chen, F. F., Hayes, A., Carver, C. S., Laurenceau, J.-P. and Zhang, Z. [2012], ‘Modeling general and specific variance in multifaceted constructs: A comparison of the bifactor model to other approaches’, Journal of personality 80(1), 219–251.

19. Chen, P.-Y., Yang, C.-M. and Morin, C. M. [2015], ‘Validating the cross-cultural factor structure and invariance property of the insomnia severity index: evidence based on ordinal EFA and CFA’, Sleep medicine 16(5), 598–603.

20. Clauw, D. J. [2015], Fibromyalgia and related conditions, in ‘Mayo Clinic Proceedings’, Vol. 90, Elsevier, pp. 680–692.

21. Clauw, D. J., Essex, M. N., Pitman, V. and Jones, K. D. [2019], ‘Reframing chronic pain as a disease, not a symptom: rationale and implications for pain management’, Postgraduate medicine 131(3), 185–198.

22. Csardi, G., Nepusz, T. et al. [2006], ‘The igraph software package for complex network research’, InterJournal, complex systems 1695(5), 1–9.

23. de Leeuw, C. A., Mooij, J. M., Heskes, T. and Posthuma, D. [2015], ‘Magma: generalized gene-set analysis of GWAS data’, PLoS computational biology 11(4), e1004219.

24. Din, H. M. and Minhatb, H. S. [2021], ‘Psychometric properties of anxiety about aging scale among Malaysian youths’, Journal of Applied Structural Equation Modeling 5(1), 9–18.

25. Dutta, D., Brummett, C. M., Moser, S. E., Fritsche, L. G., Tsodikov, A., Lee, S., Clauw, D. J. and Scott, L. J. [2020], ‘Heritability of the fibromyalgia phenotype varies by age’, Arthritis & Rheumatology 72(5), 815–823.

26. Eccleston, C., Wastell, S., Crombez, G. and Jordan, A. [2008], ‘Adolescent social development and chronic pain’, European Journal of Pain 12(6), 765–774.

27. Edelmayer, R. M., Vanderah, T. W., Majuta, L., Zhang, E.-T., Fioravanti, B., De Felice, M., Chichorro, J. G., Ossipov, M. H., King, T., Lai, J. et al. [2009], ‘Medullary pain facilitating neurons mediate allodynia in headache-related pain’, Annals of Neurology: Official Journal of the American Neurological Association and the Child Neurology Society 65(2), 184–193.

28. Elliott, K. S., Chapman, K., Day-Williams, A., Panoutsopoulou, K., Southam, L., Lindgren, C. M., Arden, N., Aslam, N., Birrell, F., Carluke, I. et al. [2013], ‘Evaluation of the genetic overlap between osteoarthritis with body mass index and height using genome-wide association scan data’, Annals of the rheumatic diseases 72(6), 935–941.

29. Eppstein, D., Löoffler, M. and Strash, D. [2010], Listing all maximal cliques in sparse graphs in near-optimal time, in ‘International Symposium on Algorithms and Computation’, Springer, pp. 403–414.

30. Fillingim, R. B., Loeser, J. D., Baron, R. and Edwards, R. R. [2016], ‘Assessment of chronic pain: domains, methods, and mechanisms’, The journal of pain 17(9), T10–T20.

31. Finnerup, N. B., Attal, N., Haroutounian, S., McNicol, E., Baron, R., Dworkin, R. H., Gilron, I., Haanpöaöa, M., Hansson, P., Jensen, T. S. et al. [2015], ‘Pharmacotherapy for neuropathic pain in adults: a systematic review and meta-analysis’, The Lancet Neurology 14(2), 162–173.

32. Firth, D. [1993], ‘Bias reduction of maximum likelihood estimates’, Biometrika 80(1), 27–38.

33. Fokkema, M. and Greiff, S. [2017], How performing PCA and CFA on the same data equals trouble.

34. Foote, I. F., Jacobs, B. M., Mathlin, G., Watson, C. J., Bothongo, P. L., Waters, S., Dobson, R., Noyce, A. J., Bhui, K. S., Korszun, A. et al. [2021], The genetic architecture of Alzheimer’s disease risk: A genomic structural equation modelling study, medRxiv.

35. Freidin, M. B., Tsepilov, Y. A., Palmer, M., Karssen, L. C., Suri, P., Aulchenko, Y. S., Williams, F. M., Group, C. M. W. et al. [2019], ‘Insight into the genetic architecture of back pain and its risk factors from a study of 509,000 individuals’, Pain 160(6), 1361.

36. Friedman, N. P., Banich, M. T. and Keller, M. C. [2021], ‘Twin studies to GWAS: there and back again’,_Trends in Cognitive Sciences.

37. Geha, P. and Waxman, S. G. [2016], ‘Pain perception: multiple matrices or one?’, JAMA neurology 73(6), 628–630.

38. Gerbing, D. W. and Hamilton, J. G. [1996], ‘Viability of exploratory factor analysis as a precursor to confirmatory factor analysis’, Structural Equation Modeling: A Multidisciplinary Journal 3(1), 62–72.

39. Gormley, P., Anttila, V., Winsvold, B. S., Palta, P., Esko, T., Pers, T. H., Farh, K.-H., Cuenca-Leon, E., Muona, M., Furlotte, N. A. et al. [2016], ‘Meta-analysis of 375,000 individuals identifies 38 susceptibility loci for migraine’, Nature genetics 48(8), 856–866.

40. Grace, P. M., Hutchinson, M. R., Maier, S. F. and Watkins, L. R. [2014], ‘Pathological pain and the neuroimmune interface’, Nature Reviews Immunology 14(4), 217–231.

41. Gregory, N. S., Harris, A. L., Robinson, C. R., Dougherty, P. M., Fuchs, P. N. and Sluka, K. A. [2013], ‘An overview of animal models of pain: disease models and outcome measures’, The Journal of Pain 14(11), 1255–1269.

42. Grotzinger, A. D., Mallard, T. T., Akingbuwa, W. A., Ip, H. F., Adams, M. J., Lewis, C. M., McIntosh, A. M., Grove, J., Dalsgaard, S., Peter-Lesch, K. et al. [2020], ‘Genetic architecture of 11 major psychiatric disorders at biobehavioral, functional genomic, and molecular genetic levels of analysis’, medRxiv.

43. Grotzinger, A. D., Rhemtulla, M., de Vlaming, R., Ritchie, S. J., Mallard, T. T., Hill, W. D., Ip, H. F., Marioni, R. E., McIntosh, A. M., Deary, I. J. et al. [2019], ‘Genomic structural equation modelling provides insights into the multivariate genetic architecture of complex traits’, Nature human behaviour 3(5), 513–525.

44. Han, X., Ashar, Y. K., Kragel, P., Petre, B., Schelkun, V., Atlas, L. Y., Chang, L. J., Jepma, M., Koban, L., Losin, E. A. R. et al. [2022], ‘Effect sizes and test-retest reliability of the fMRI-based neurologic pain signature’, NeuroImage 247, 118844.

45. Hartvigsen, J., Nielsen, J., Kyvik, K. O., Fejer, R., Vach, W., Iachine, I. and Leboeuf-Yde, C. [2009], ‘Heritability of spinal pain and consequences of spinal pain: A comprehensive genetic epidemiologic analysis using a population-based sample of 15,328 twins ages 20-71 years’, Arthritis Care & Research 61(10), 1343–1351.

46. Hengstebeck, E., Roskos, S., Breejen, K., Arnetz, B. and Arnetz, J. [2017], ‘Chronic pain disrupts ability to work by interfering with social function: a cross-sectional study’, Scandinavian journal of pain 17(1), 397–402.

47. Howard, M. A., Krause, K., Khawaja, N., Massat, N., Zelaya, F., Schumann, G., Huggins, J. P., Vennart, W., Williams, S. C. and Renton, T. F. [2011], ‘Beyond patient reported pain: perfusion magnetic resonance imaging demonstrates reproducible cerebral representation of ongoing post-surgical pain’, PloS One 6(2), e17096.

48. Hruschak, V. and Cochran, G. [2018], ‘Psychosocial predictors in the transition from acute to chronic pain: a systematic review’, Psychology, health & medicine 23(10), 1151–1167.

49. Hu, L.-t. and Bentler, P. M. [1999], ‘Cutoff criteria for fit indexes in covariance structure analysis: Conventional criteria versus new alternatives’, Structural equation modeling: a multidisciplinary journal 6(1), 1–55.

50. Huang, J., Howie, B., McCarthy, S., Memari, Y., Walter, K., Min, J. L., Danecek, P., Malerba, G., Trabetti, E., Zheng, H.-F. et al. [2015], ‘Improved imputation of low-frequency and rare variants using the UK10K haplotype reference panel’, Nature communications 6(1), 1–9.

51. Huedo-Medina, T. B., S’ sanchez-Meca, J., Marin-Martinez, F. and Botella, J. [2006], ‘Assessing heterogeneity in meta-analysis: Q statistic or I^2^ index?’, Psychological methods 11(2), 193.

52. Jefferson, T., Kelly, C. J. and Martina, M. [2021], ‘Differential rearrangement of excitatory inputs to the medial prefrontal cortex in chronic pain models’, Frontiers in Neural Circuits p. 159.

53. Jensen, A. R. [2000], ‘The g factor: psychometrics and biology’, The nature of intelligence p. 37.

54. Ji, R.-R., Nackley, A., Huh, Y., Terrando, N. and Maixner, W. [2018], ‘Neuroinflammation and central sensitization in chronic and widespread pain’, Anesthesiology 129(2), 343–366.

55. Ji, R.-R., Xu, Z.-Z. and Gao, Y.-J. [2014], ‘Emerging targets in neuroinflammation-driven chronic pain’, Nature reviews Drug discovery 13(7), 533–548.

56. Johnston, K. J., Adams, M. J., Nicholl, B. I., Ward, J., Strawbridge, R. J., Ferguson, A., McIntosh, \A. M., Bailey, M. E. and Smith, D. J. [2019], ‘Genome-wide association study of multisite chronic pain in UK Biobank’, PLoS genetics 15(6), e1008164.

57. Johnston, K. J., Ward, J., Ray, P. R., Adams, M. J., McIntosh, A. M., Smith, B. H., Strawbridge, R. J., Price, T. J., Smith, D. J., Nicholl, B. I. et al. [2021], ‘Sex-stratified genome-wide association study of multisite chronic pain in UK Biobank’, PLoS genetics 17(4), e1009428.

58. Kambur, O., Kaunisto, M. A., Winsvold, B. S., Wilsgaard, T., Stubhaug, A., Zwart, J. A., Kalso, E. and Nielsen, C. S. [2018], ‘Genetic variation in P2RX7 and pain tolerance’, Pain 159(6), 1064.

59. Khoury, S., Parisien, M., Thompson, S. J., Vachon-Presseau, E., Roy, M., Martinsen, A. E., Winsvold, B. S., Pain, H. A.-I., Mundal, I. P., Zwart, J.-A. et al. [2021], ‘Genome-wide analysis identifies impaired axonogenesis in chronic overlapping pain conditions’, Brain.

60. Kline, R. B. [2015], Principles and practice of structural equation modeling, Guilford publications.

61. Kong, J., Spaeth, B., Wey, H.-Y., Cheetham, A., Cook, A. H., Jensen, K., Tan, Y., Liu, H., Wang, D., Loggia, M. L. et al. [2013], ‘S1 is associated with chronic low back pain: a functional and structural MRI study’, Molecular pain 9, 1744–8069.

62. Kucyi, A. and Davis, K. D. [2015], ‘The dynamic pain connectome’, Trends in neurosciences 38(2), 86–95.

63. Kuner, R. and Flor, H. [2017], ‘Structural plasticity and reorganisation in chronic pain’, Nature Reviews Neuroscience 18(1), 20–30.

64. Kyriazos, T. A. et al. [2018], ‘Applied psychometrics: sample size and sample power considerations in factor analysis (EFA, CFA) and SEM in general’, Psychology 9(08), 2207.

65. Lacagnina, M. J., Heijnen, C. J., Watkins, L. R. and Grace, P. M. [2021], ‘Autoimmune regulation of chronic pain’, Pain reports 6(1).

66. Lau, C.-I., Lin, C.-C., Chen, W.-H., Wang, H.-C. and Kao, C.-H. [2014], ‘Association between migraine and irritable bowel syndrome: A population-based retrospective cohort study’, European Journal of Neurology 21(9), 1198–1204.

67. Lee, J.-J., Kim, H. J., Cěko, M., Park, B.-y., Lee, S. A., Park, H., Roy, M., Kim, S.-G., Wager, T. D. and Woo, C.-W. [2021], ‘A neuroimaging biomarker for sustained experimental and clinical pain’, Nature medicine 27(1), 174–182.

68. Lee, J., Protsenko, E., Lazaridou, A., Franceschelli, O., Ellingsen, D.-M., Mawla, I., Isenburg, K., Berry, M. P., Galenkamp, L., Loggia, M. L. et al. [2018], ‘Encoding of self-referential pain catastrophizing in the posterior cingulate cortex in fibromyalgia’, Arthritis & rheumatology 70(8), 1308–1318.

69. Lee, S. H., Wray, N. R., Goddard, M. E. and Visscher, P. M. [2011], ‘Estimating missing heritability for disease from genome-wide association studies’, The American Journal of Human Genetics 88(3), 294–305.

70. Loggia, M. L., Berna, C., Kim, J., Cahalan, C. M., Martel, M.-O., Gollub, R. L., Wasan, A. D., Napadow, V. and Edwards, R. R. [2015], ‘The lateral prefrontal cortex mediates the hyperalgesic effects of negative cognitions in chronic pain patients’, The Journal of Pain 16(8), 692–699.

71. Lonsdale, J., Thomas, J., Salvatore, M., Phillips, R., Lo, E., Shad, S., Hasz, R., Walters, G., Garcia, F., Young, N. et al. [2013], ‘The genotype-tissue expression (GTEx) project’, Nature genetics 45(6), 580–585.

72. Maixner, W., Fillingim, R. B., Williams, D. A., Smith, S. B. and Slade, G. D. [2016], ‘Overlapping chronic pain conditions: implications for diagnosis and classification’, The Journal of Pain 17(9), T93–T107.

73. Malkin, I., Williams, F. M., LaChance, G., Spector, T., MacGregor, A. J. and Livshits, G. [2014], ‘Low back and common widespread pain share common genetic determinants’, Annals of human genetics 78(5), 357–366.

74. Mallard, T. T., Linn’ ser, R. K., Grotzinger, A. D., Sanchez-Roige, S., Seidlitz, J., Okbay, A., de Vlaming, R., Meddens, S. F. W., Palmer, A. A., Davis, L. K. et al. [2020], ‘Multivariate GWAS of psychiatric disorders and their cardinal symptoms reveal two dimensions of cross-cutting genetic liabilities’, BioRxiv, p. 603134.

75. Mansour, A., Farmer, M., Baliki, M. and Apkarian, A. V. [2014], ‘Chronic pain: the role of learning and brain plasticity’, Restorative neurology and neuroscience 32(1), 129–139.

76. Martins, D., Dipasquale, O., Veronese, M., Turkheimer, F., Loggia, M. L., McMahon, S., Howard, M. A. and Williams, S. C. [2021], ‘Transcriptional and cellular signatures of cortical morphometric remodelling in chronic pain’, Pain.

77. Mayer, S., Spickschen, J., Stein, K. V., Crevenna, R., Dorner, T. E. and Simon, J. [2019], ‘The societal costs of chronic pain and its determinants: the case of Austria’, PloS One 14(3), e0213889.

78. Mbatchou, J., Barnard, L., Backman, J., Marcketta, A., Kosmicki, J. A., Ziyatdinov, A., Benner, C., O’Dushlaine, C., Barber, M., Boutkov, B. et al. [2020], ‘Computationally efficient whole genome regression for quantitative and binary traits’, bioRxiv.

79. Meints, S. M., Mawla, I., Napadow, V., Kong, J., Gerber, J., Chan, S.-T., Wasan, A. D., Kaptchuk, T. J., McDonnell, C., Carriere, J. et al. [2019], ‘The relationship between catastrophizing and altered pain sensitivity in patients with chronic low back pain’, Pain 160(4), 833.

80. Melzack, R. [2001], ‘Pain and the neuromatrix in the brain’, Journal of dental education 65(12), 1378–1382.

81. Meng, W., Adams, M. J., Hebert, H. L., Deary, I. J., McIntosh, A. M. and Smith, B. H. [2018], ‘A genome-wide association study finds genetic associations with broadly-defined headache in UK Biobank (n= 223,773)’, EBioMedicine 28, 180–186.

82. Meng, W., Adams, M. J., Palmer, C. N., Shi, J., Auton, A., Ryan, K. A., Jordan, J. M., Mitchell, B. D., Jackson, R. D., Yau, M. S. et al. [2019], ‘Genome-wide association study of knee pain identifies associations with GDF5 and COL27A1 in UK Biobank’, Communications biology 2(1), 1–8.

83. Meng, W., Adams, M. J., Reel, P., Rajendrakumar, A., Huang, Y., Deary, I. J., Palmer, C. N., McIntosh, A. M. and Smith, B. H. [2020], ‘Genetic correlations between pain phenotypes and depression and neuroticism’, European Journal of Human Genetics 28(3), 358–366.

84. Meng, W., Chan, B. W., Harris, C., Freidin, M. B., Hebert, H. L., Adams, M. J., Campbell, A., Hayward, C., Zheng, H., Zhang, X. et al. [2020], ‘A genome-wide association study finds genetic variants associated with neck or shoulder pain in UK Biobank’, Human Molecular Genetics 29(8), 1396–1404.

85. Meng, W., Deshmukh, H., Van Zuydam, N., Liu, Y., Donnelly, L., Zhou, K., Wellcome Trust Case Control Consortium 2 (WTCCC2), Surrogate Markers for Micro- and Macro-Vascular Hard Endpoints for Innovative Diabetes Tools (SUMMIT) Study Group, S. M., Morris, A., Colhoun, H.M., Palmer, C.N.A., Smith, B.H. [2015], A genome-wide association study suggests an association of Chr8p21. 3 (GFRA2) with diabetic neuropathic pain, European Journal of Pain 19(3), 392–399.

86. Meng, W., Reel, P., Nangia, C., Rajendrakumar, A., Hebert, H., Adams, M., Zheng, H., Lu, Z., Ray, D., Colvin, L. et al. [2021], ‘A meta-analysis of the genome-wide association studies on two genetically correlated phenotypes (self-reported headache and self-reported migraine) identifies four new risk loci for headaches (n= 397,385)’, medRxiv.

87. Miller Neilan, R., Majetic, G., Gil-Silva, M., Adke, A. P., Carrasquillo, Y. and Kolber, B. J. [2021], ‘Agent-based modeling of the central amygdala and pain using cell-type specific physiological parameters’, PLoS Computational Biology 17(6), e1009097.

88. Miyake, A., Friedman, N. P., Emerson, M. J., Witzki, A. H., Howerter, A. and Wager, T. D. [2000], ‘The unity and diversity of executive functions and their contributions to complex frontal lobe tasks: A latent variable analysis’, Cognitive psychology 41(1), 49–100.

89. Morlion, B., Coluzzi, F., Aldington, D., Kocot-Kepska, M., Pergolizzi, J., Mangas, A. C., Ahlbeck, K. and Kalso, E. [2018], ‘Pain chronification: what should a non-pain medicine specialist know?’, Current Medical Research and Opinion 34(7), 1169–1178.

90. Napadow, V. and Harris, R. E. [2014], ‘What has functional connectivity and chemical neuroimaging in fibromyalgia taught us about the mechanisms and management of centralized’pain?’, Arthritis research & therapy 16(4), 1–8.

91. Navratilova, E. and Porreca, F. [2014], ‘Reward and motivation in pain and pain relief’, Nature neuroscience 17(10), 1304–1312.

92. Nicholas, M., Vlaeyen, J. W., Rief, W., Barke, A., Aziz, Q., Benoliel, R., Cohen, M., Evers, S., Giamberardino, M. A., Goebel, A. et al. [2019], ‘The IASP classification of chronic pain for icd-11: chronic primary pain’, Pain 160(1), 28–37.

93. Niv, D. and Devor, M. [2004], ‘Chronic pain as a disease in its own right’. Pain Practice 4.3 (2004): 179–181.

94. O’Connor, L. J., Schoech, A. P., Hormozdiari, F., Gazal, S., Patterson, N. and Price, A. L. [2019], ‘Extreme polygenicity of complex traits is explained by negative selection’, The American Journal of Human Genetics 105(3), 456–476.

95. Osborne, N. R., Anastakis, D. J. and Davis, K. D. [2018], ‘Peripheral nerve injuries, pain, and neuro-plasticity’, Journal of Hand Therapy 31(2), 184–194.

96. Ossipov, M. H., Morimura, K. and Porreca, F. [2014], ‘Descending pain modulation and chronification of pain’, Current opinion in supportive and palliative care 8(2), 143.

97. Phillips, C. J. [2006], ‘Economic burden of chronic pain’, Expert review of pharmacoeconomics & out-comes research 6(5), 591–601.

98. Plesh, O., Noonan, C., Buchwald, D. S., Goldberg, J. and Afari, N. [2012], ‘Temporomandibular disorder-type pain and migraine headache in women: a preliminary twin study’, Journal of orofacial pain 26(2), 91.

99. Price, A. L., Patterson, N. J., Plenge, R. M., Weinblatt, M. E., Shadick, N. A. and Reich, D. [2006], ‘Principal components analysis corrects for stratification in genome-wide association studies’, Nature genetics 38(8), 904–909.

100. Quesada, C., Kostenko, A., Ho, I., Leone, C., Nochi, Z., Stouffs, A., Wittayer, M., Caspani, O., Finnerup, N. B., Mouraux, A. et al. [2021], ‘Human surrogate models of central sensitization: A critical review and practical guide’, European journal of pain (London, England) 25(7), 1389.

101. Raffaeli, W. and Arnaudo, E. [2017], ‘Pain as a disease: an overview’, Journal of pain research 10, 2003.

102. Sakaue, S., Kanai, M., Tanigawa, Y., Karjalainen, J., Kurki, M., Koshiba, S., Narita, A., Konuma, T., Yamamoto, K., Akiyama, M. et al. [2021], ‘A cross-population atlas of genetic associations for 220 human phenotypes’, Nature genetics 53(10), 1415–1424.

103. Schubert, A.-L., Held, M., Sommer, C. and Üçeyler, N. [2019], ‘Reduced gene expression of netrin family members in skin and sural nerve specimens of patients with painful peripheral neuropathies’, Journal of neurology 266(11), 2812–2820.

104. Schug, S. A. and Bruce, J. [2017], ‘Risk stratification for the development of chronic postsurgical pain’, Pain Reports 2(6).

105. Schur, E. A., Afari, N., Furberg, H., Olarte, M., Goldberg, J., Sullivan, P. F. and Buchwald, D. [2007], ‘Feeling bad in more ways than one: comorbidity patterns of medically unexplained and psychiatric conditions’, Journal of general internal medicine 22(6), 818–821.

106. Seminowicz, D. A., Labus, J. S., Bueller, J. A., Tillisch, K., Naliboff, B. D., Bushnell, M. C. and Mayer, E. A. [2010], ‘Regional gray matter density changes in brains of patients with irritable bowel syndrome’, Gastroenterology 139(1), 48–57.

107. Sibille, K. T., Steingrímsdöttir, Ó. A., Fillingim, R. B., Stubhaug, A., Schirmer, H., Chen, H., McEwen, B. S. and Nielsen, C. S. [2016], ‘Investigating the burden of chronic pain: an inflammatory and metabolic composite’, Pain Research and Management 2016.

108. Simon, L. S. [2012], ‘Relieving pain in America: A blueprint for transforming prevention, care, education, and research’, Journal of pain & palliative care pharmacotherapy 26(2), 197–198.

109. Smith, S. M., Dworkin, R. H., Turk, D. C., Baron, R., Polydefkis, M., Tracey, I., Borsook, D., Edwards, R. R., Harris, R. E., Wager, T. D. et al. [2017], ‘The potential role of sensory testing, skin biopsy, and functional brain imaging as biomarkers in chronic pain clinical trials: Impact considerations’, The Journal of Pain 18(7), 757–777.

110. Sommer, C., Leinders, M. and Üçeyler, N. [2018], ‘Inflammation in the pathophysiology of neuropathic pain’, Pain 159(3), 595–602.

111. Stanos, S., Brodsky, M., Argoff, C., Clauw, D. J., DArcy, Y., Donevan, S., Gebke, K. B., Jensen, M. P., Lewis&Clark, E., McCarberg, B. et al. [2016], ‘Rethinking chronic pain in a primary care setting’, Postgraduate Medicine 128(5), 502–515.

112. Supek, F., Bošnjak, M., Škunca, N. and Šmuc, T. [2011], ‘Revigo summarizes and visualizes long lists of gene ontology terms’, PloS one 6(7), e21800.

113. Suri, P., Morgenroth, D. C., Kwoh, C. K., Bean, J. F., Kalichman, L. and Hunter, D. J. [2010], ‘Low back pain and other musculoskeletal pain comorbidities in individuals with symptomatic osteoarthritis of the knee: data from the osteoarthritis initiative’, Arthritis care & research 62(12), 1715–1723.

114. Suri, P., Palmer, M. R., Tsepilov, Y. A., Freidin, M. B., Boer, C. G., Yau, M. S., Evans, D. S., Gelemanovic, A., Bartz, T. M., Nethander, M. et al. [2018], ‘Genome-wide meta-analysis of 158,000 individuals of European ancestry identifies three loci associated with chronic back pain’, PLoS genetics 14(9), e1007601.

115. Tian, C., Gregersen, P. K. and Seldin, M. F. [2008], ‘Accounting for ancestry: population substructure and genome-wide association studies’, Human molecular genetics 17(R2), R143–R150.

116. Tracey, I. and Bushnell, M. C. [2009], ‘How neuroimaging studies have challenged us to rethink: is chronic pain a disease?’, The journal of pain 10(11), 1113–1120.

117. Treede, R.-D., Kenshalo, D. R., Gracely, R. H. and Jones, A. K. [1999], ‘The cortical representation of pain’, Pain 79(2-3), 105–111.

118. Treede, R.-D., Rief, W., Barke, A., Aziz, Q., Bennett, M. I., Benoliel, R., Cohen, M., Evers, S., Finnerup, N. B., First, M. B. et al. [2015], ‘A classification of chronic pain for icd-11’, Pain 156(6), 1003.

119. Treede, R.-D., Rief, W., Barke, A., Aziz, Q., Bennett, M. I., Benoliel, R., Cohen, M., Evers, S., Finnerup, N. B., First, M. B. et al. [2019], ‘Chronic pain as a symptom or a disease: the IASP classification of chronic pain for the international classification of diseases (icd-11)’, Pain 160(1), 19–27.

120. Tsepilov, Y. A., Freidin, M. B., Shadrina, A. S., Sharapov, S. Z., Elgaeva, E. E., van Zundert, J., Karssen, L., Suri, P., Williams, F. M. and Aulchenko, Y. S. [2020], ‘Analysis of genetically independent phenotypes identifies shared genetic factors associated with chronic musculoskeletal pain conditions’, Communications biology 3(1), 1–13.

121. Tylee, D. S., Sun, J., Hess, J. L., Tahir, M. A., Sharma, E., Malik, R., Worrall, B. B., Levine, A. J., Martinson, J. J., Nejentsev, S. et al. [2018], ‘Genetic correlations among psychiatric and immune-related phenotypes based on genome-wide association data’, American Journal of Medical Genetics Part B: Neuropsychiatric Genetics 177(7), 641–657.

122. Urien, L. and Wang, J. [2019], ‘Top-down cortical control of acute and chronic pain’, Psychosomatic medicine 81(9), 851.

123. Vachon-Presseau, E., Berger, S. E., Abdullah, T. B., Griffith, J. W., Schnitzer, T. J. and Apkarian, A. V. [2019], ‘Identification of traits and functional connectivity-based neurotraits of chronic pain’, PLoS biology 17(8), e3000349.

124. Vachon-Presseau, E., Centeno, M., Ren, W., Berger, S., Tétreault, P., Ghantous, M., Baria, A., Farmer, M., Baliki, M., Schnitzer, T. et al. [2016], ‘The emotional brain as a predictor and amplifier of chronic pain’, Journal of dental research 95(6), 605–612.

125. Vachon-Presseau, E., Roy, M., Martel, M.-O., Caron, E., Marin, M.-F., Chen, J., Albouy, G., Plante, I., Sullivan, M. J., Lupien, S. J. et al. [2013], ‘The stress model of chronic pain: evidence from basal cortisol and hippocampal structure and function in humans’, Brain 136(3), 815–827.

126. Vachon-Presseau, E., T’ setreault, P., Petre, B., Huang, L., Berger, S. E., Torbey, S., Baria, A. T., Mansour, A. R., Hashmi, J. A., Griffith, J. W. et al. [2016], ‘Corticolimbic anatomical characteristics predetermine risk for chronic pain’, Brain 139(7), 1958–1970.

127. van Bork, R., Epskamp, S., Rhemtulla, M., Borsboom, D. and van der Maas, H. L. [2017], ‘What is the p-factor of psychopathology? some risks of general factor modeling’, Theory & Psychology 27(6), 759–773.

128. van der Gaag, W. H., Roelofs, P. D., Enthoven, W. T., van Tulder, M. W. and Koes, B. W. [2020], ‘Non-steroidal anti-inflammatory drugs for acute low back pain’, Cochrane Database of Systematic Reviews (4).

129. van Reij, R. R., Voncken, J. W., Joosten, E. A. and van den Hoogen, N. J. [2020], ‘Polygenic risk scores indicates genetic overlap between peripheral pain syndromes and chronic postsurgical pain’, Neurogenetics 21(3), 205–215.

130. Vehof, J., Zavos, H. M., Lachance, G., Hammond, C. J. and Williams, F. M. [2014], ‘Shared genetic factors underlie chronic pain syndromes’, PAIN® 155(8), 1562–1568.

131. Walitt, B., Ceko, M., L Gracely, J. and H Gracely, R. [2016], ‘Neuroimaging of central sensitivity syndromes: key insights from the scientific literature’, Current rheumatology reviews 12(1), 55–87.

132. Watanabe, K., Stringer, S., Frei, O., Umi’ scevi’c Mirkov, M., de Leeuw, C., Polderman, T. J., van der Sluis, S., Andreassen, O. A., Neale, B. M. and Posthuma, D. [2019], ‘A global overview of pleiotropy and genetic architecture in complex traits’, Nature genetics 51(9), 1339–1348.

133. Watanabe, K., Taskesen, E., Van Bochoven, A. and Posthuma, D. [2017], ‘Functional mapping and annotation of genetic associations with fuma’, Nature communications 8(1), 1–11.

134. WHO [2019], ‘Musculoskeletal conditions.’, https://www.who.int/news-room/fact-sheets/detail/musculoskeletal-conditions. Accessed: 2021-12-16.

135. Wiberg, A., Ng, M., Schmid, A. B., Smillie, R. W., Baskozos, G., Holmes, M. V., Kunnapuu, K., Mägi, R., Bennett, D. L. and Furniss, D. [2019], ‘A genome-wide association analysis identifies 16 novel susceptibility loci for carpal tunnel syndrome’, Nature communications 10(1), 1–12.

136. Williams, F. M., Spector, T. D. and MacGregor, A. J. [2010], ‘Pain reporting at different body sites is explained by a single underlying genetic factor’, Rheumatology 49(9), 1753–1755.

137. Woolf, C. J. [2011], ‘Central sensitization: implications for the diagnosis and treatment of pain’, pain 152(3), S2–S15.

138. Wray, N. R., Wijmenga, C., Sullivan, P. F., Yang, J. and Visscher, P. M. [2018], ‘Common disease is more complex than implied by the core gene omnigenic model’, Cell 173(7), 1573–1580.

139. Wrigley, P. J., Press, S. R., Gustin, S. M., Macefield, V. G., Gandevia, S. C., Cousins, M. J., Middleton, J. W., Henderson, L. A. and Siddall, P. [2009], ‘Neuropathic pain and primary somatosensory cortex reorganization following spinal cord injury’, Pain 141(1-2), 52–59.

