## Supplementary Note and Figures for "Identification and characterization of genetic risk shared across 24 chronic pain conditions in the UK Biobank": SupplementaryNote+Figures.pdf

##### *EFA-CFA model comparison with anatomic and etiologic models*

The EFA-CFA model with specific factor F2 encompassing musculoskeletal conditions provided a better fit than either the anatomic or the etiologic model. In the anatomic model, the Leg/Foot, Pelvic, and Torso specific factors had largely non-significant loadings from their indicators, suggesting that shared variance for these conditions was explained primarily by the general factor, F1. The 5 residual variances initially estimated to be negative also show that the model as specified is not an optimal fit for the data. The etiologic model, on the other hand, with 4 of 6 F2 conditions having non-significant loadings, shows that this factor does not explain sufficient shared variance for conditions grouped by putative inflammatory etiology beyond what is explained by the general factor.

##### *Validation of the EFA-CFA model*

To validate our findings for the EFA-CFA model, we split the genome into odd and even autosomes. Then we ran EFA on the odd set and used its output to specify loadings in Genomic SEM on the same set of chromosomes for CFA. Then we ran the same model that we used for CFA in the even chromosomes in GenomicSEM and compared the fit indices. The odd chromosome model yielded a CFI of 0.88 and SRMR of 0.12 and the even one a CFI of 0.90 and SRMR of 0.13. The factor structure for this validation model was similar to the main whole-genome model, except there were 2 specific factors: one with loadings from arthropathies, back pain, Carpal tunnel, enthesopathies of the lower limb, CWP, hip pain, hip arthrosis, knee pain, knee arthrosis, leg pain, neck pain, enthesopathies, pain in joint, and the other with loadings from back pain, chest pain (baseline), chest pain during physical activity, cystitis, gastritis, CWP, gout, oesophagitis, rheumatoid arthritis, and stomach pain. All loadings were significant, except for cystitis on the second specific factor. Given these comparable metrics in the training, validation, and whole genome datasets we concluded that using EFA and CFA on the same dataset did not result in substantial overfitting.

##### *Pathway analysis*

Starting with the list of genes mapped using all 3 approaches in FUMA – positional, eQTL, and chromatin interactions – we used GeneSCF Subhash and Kanduri (2016) to assign gene ontology (GO) pathway IDs to them and submitted the resulting list of GO IDs with the corresponding gene association $p$ values to REVIGO Supek *et al.* (2011), which reduces redundancies and aids in classification and interpretation of biological pathways. For REVIGO, we specified the options: resulting list "Small (0.5)", values associated with GO terms represent "$p$ value", remove obsolete GO terms "Yes", species "Homo sapiens (9606)", and semantic similarity measure "SimRel (default)". The resulting list cutoff 0.5 is the default similarity threshold which marks any pathways with a similarity score greater than that value as redundant, thereby shortening the list. REVIGO provides "uniqueness", which measures the extent to which a given term is an outlier when semantically compared to the complete list of submitted terms, and "dispensability", the extent to which a term is semantically close to other terms, based on semantic distance and association statistics. The term's "frequency" reflects the percentage of proteins assigned to it in the reference database (UniProt, https://www.uniprot.org/proteomes/UP000005640), such that higher frequency is an attribute of a more general term.

### Literature Cited


Subhash, S. and C. Kanduri, 2016 Genescf: a real-time based functional enrichment tool with support for multiple organisms. BMC bioinformatics **17**: 1–10.

Supek, F., M. Bošnjak, N. Škunca, and T. Šmuc, 2011 Revigo summarizes and visualizes long lists of gene ontology terms. PloS one **6**: e21800.

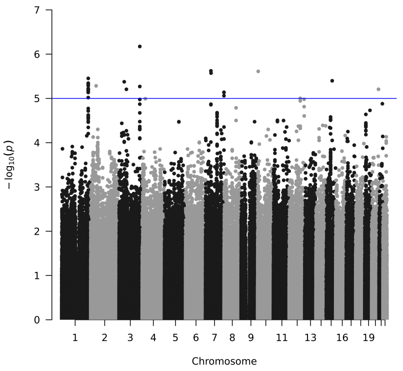


**(a)** aCMC

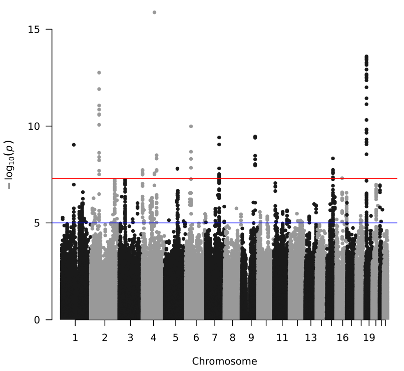


**(b)** arth

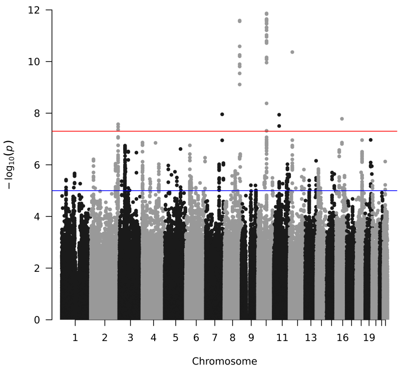


**(c)** back

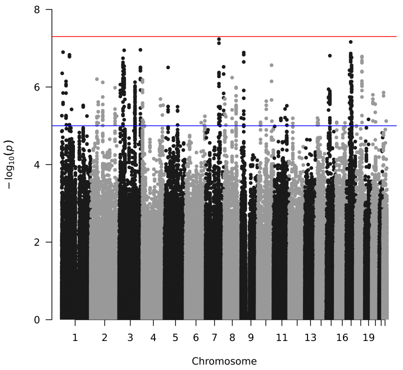


**(d)** chDs

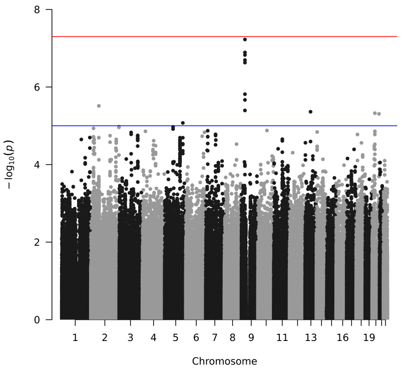


**(e)** chPh

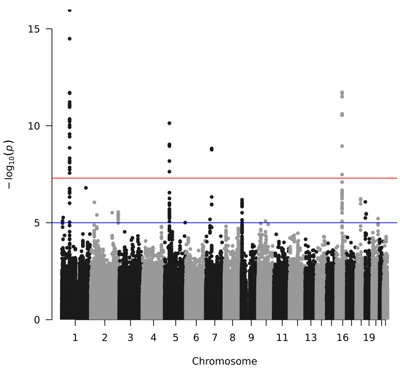


**(f)** Crhn

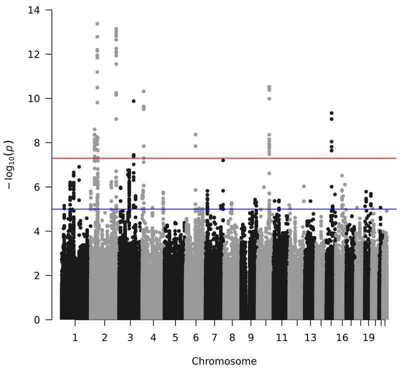


**(g)** crpl

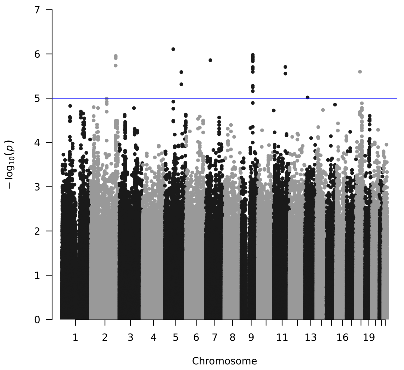


**(h)** CWP

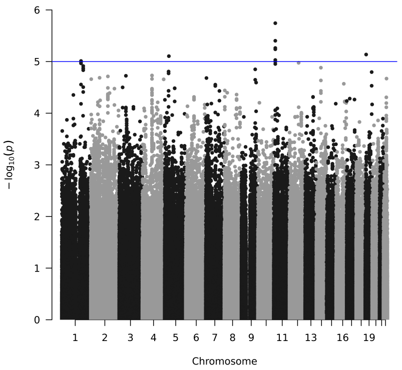


**(i)** cyst

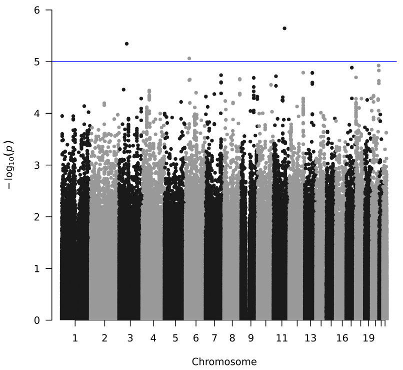


**(j)** dbNr

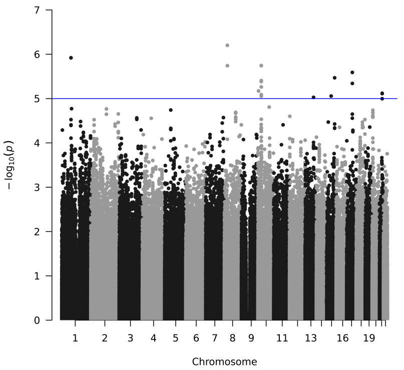


**(k)** enLL

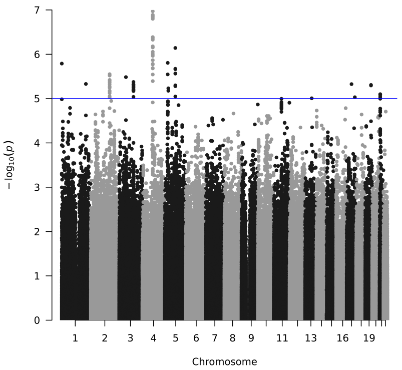


**(l)** enth

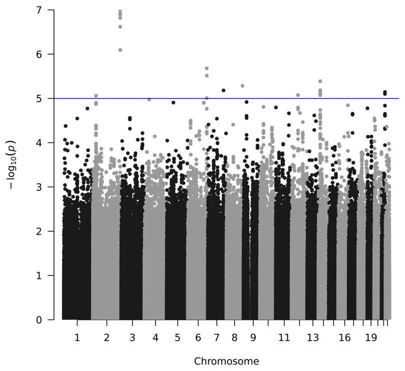


**(m)** FM

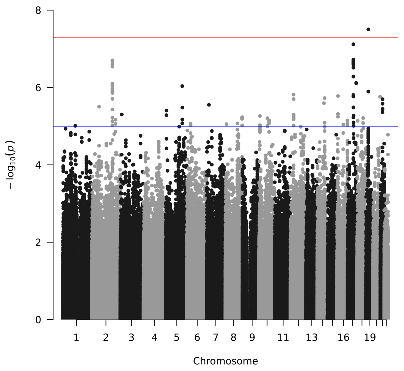


**(n)** gast

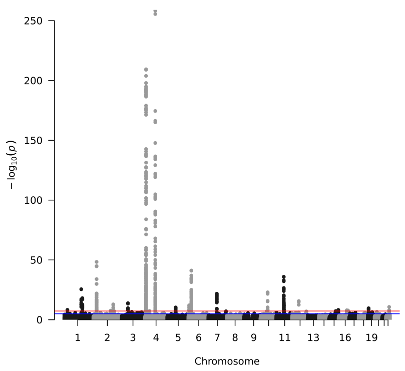


**(o)** gout

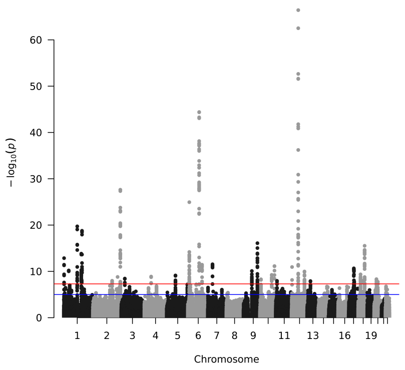


**(p)** hdch

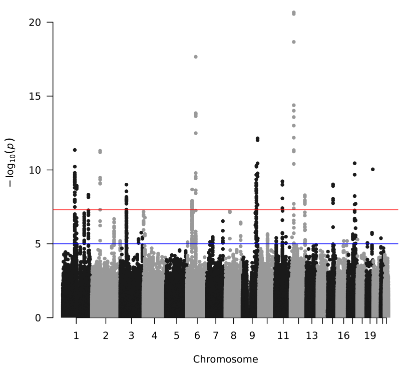


**(q)** hipA

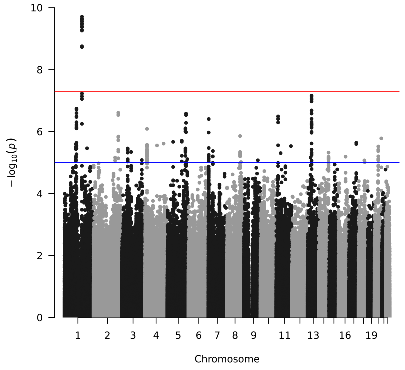


**(r)** hipP

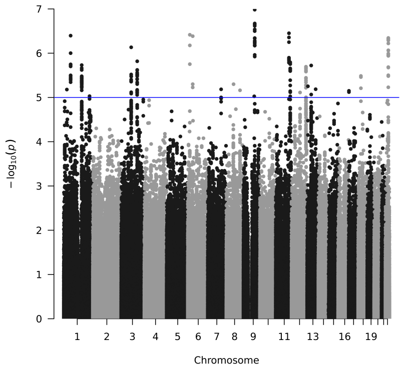


**(s)** IBS

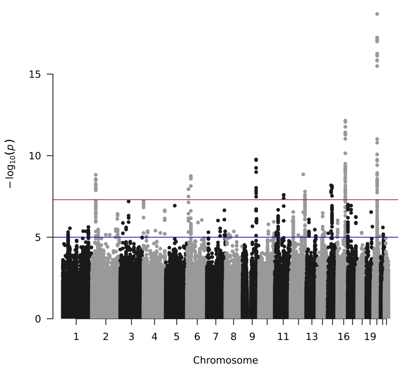


**(t)** kneA

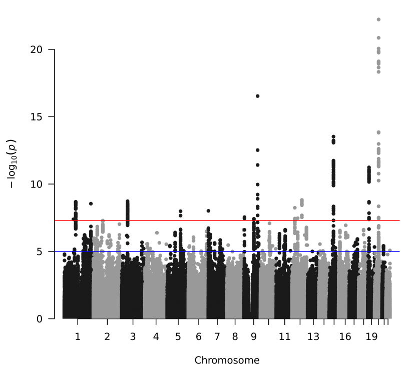


**(u)** kneP

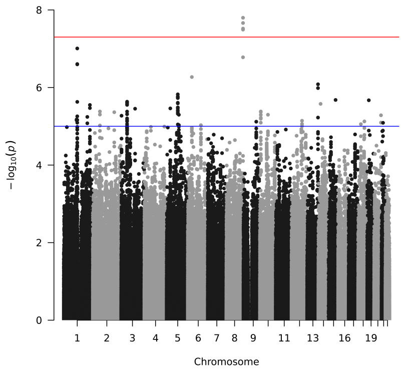


**(v)** legP

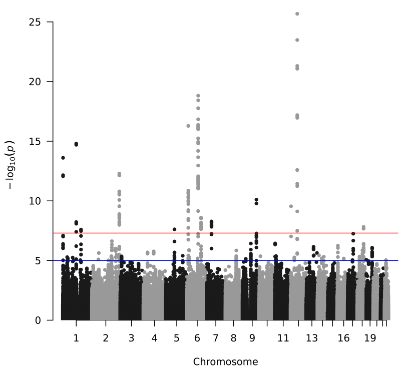


**(w)** mgrn

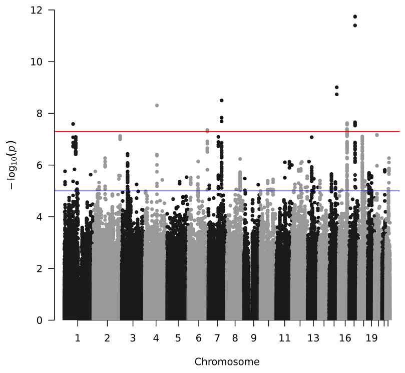


**(x)** neck

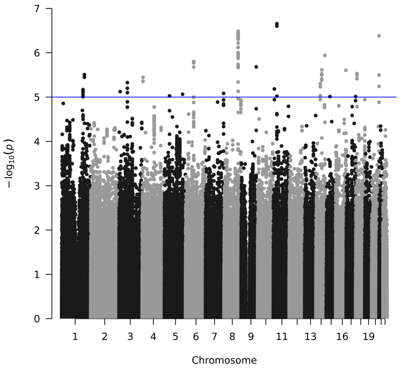


**(y)** oesp

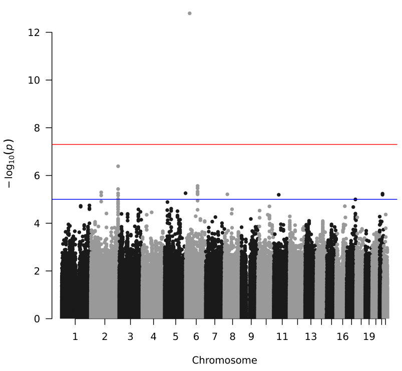


**(z)** plrh

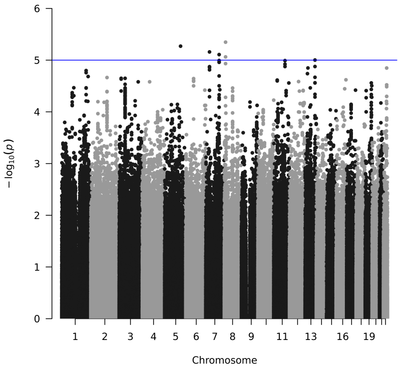


**(aa)** pnjt

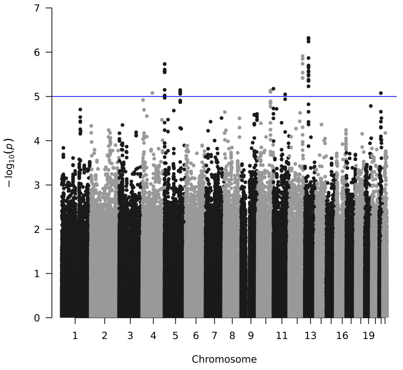


**(ab)** prst

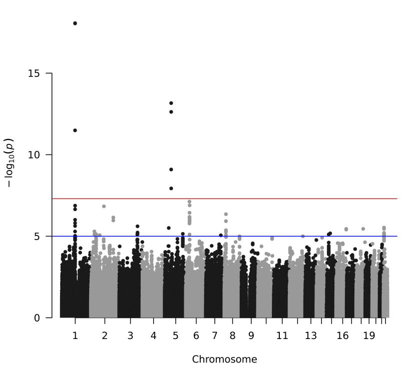


**(ac)** rhAt

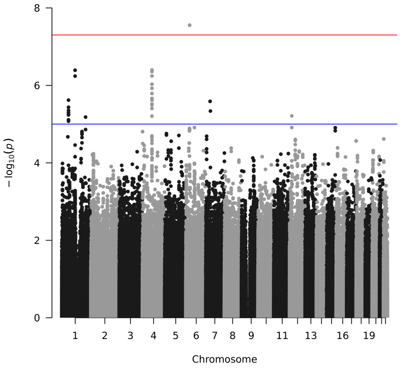


**(ad)** seRA

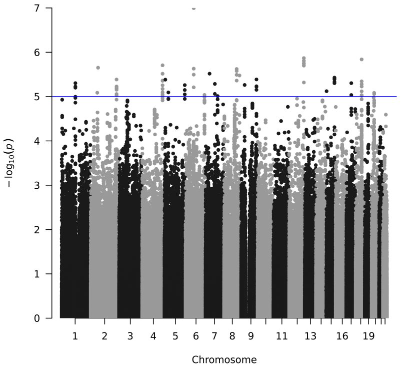


**(ae)** stmP

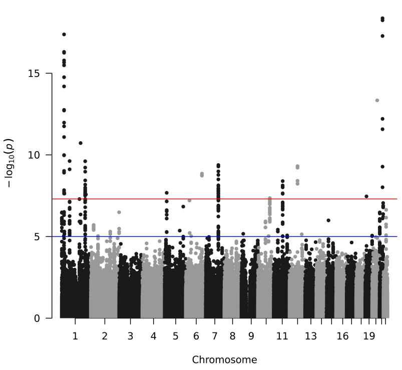


**(af)** ulcC

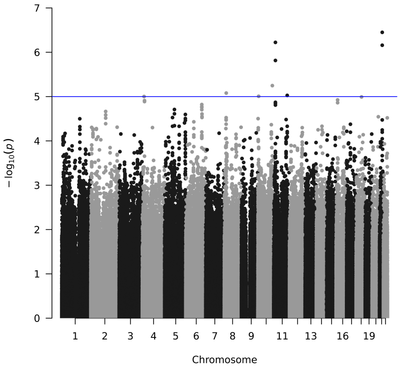


**(ag)** urCl

**Figure S1** Manhattan plots for 33 pain conditions. Condition definitions are in Table 1, and details are in Supplementary Table 1 ).

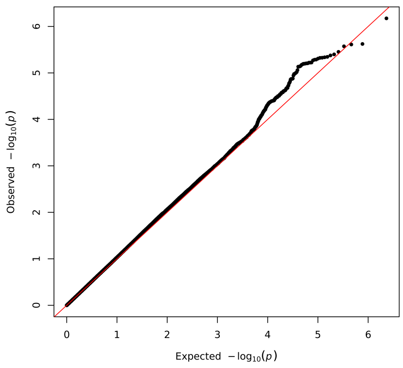


**(a)** aCMC

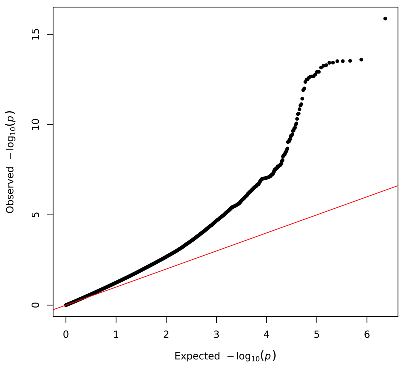


**(b)** arth

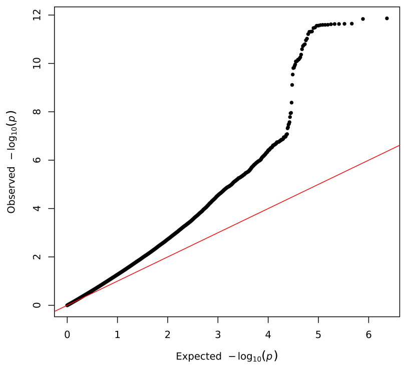


**(c)** back

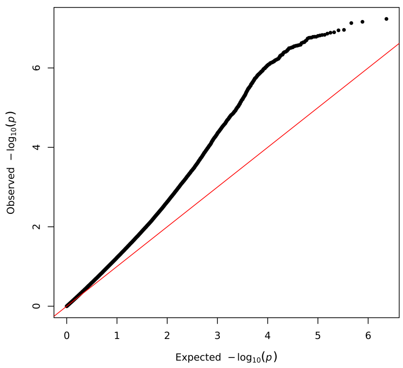


**(d)** chDs

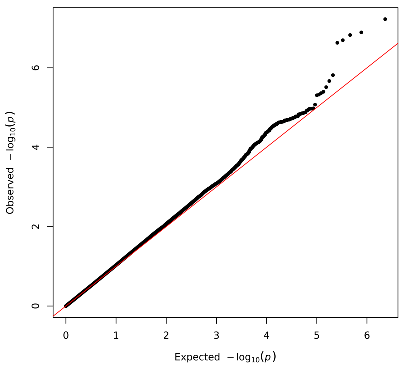


**(e)** chPh

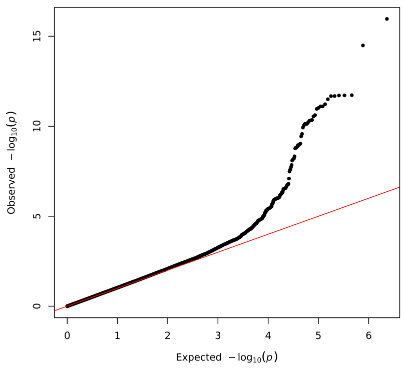


**(f)** Crhn

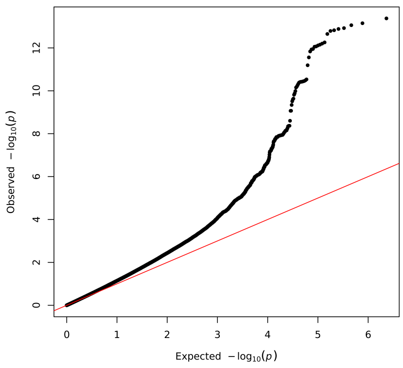


**(g)** crpl

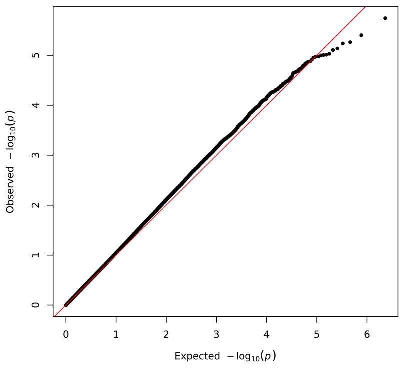


**(h)** cyst

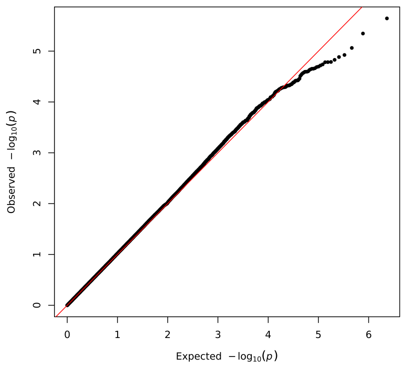


**(i)** dbNr

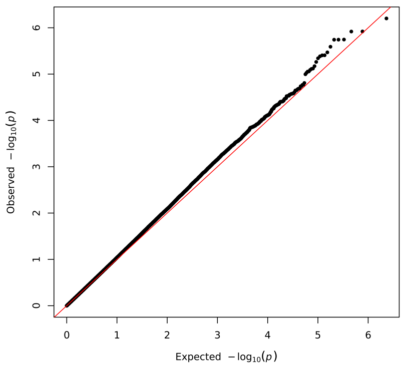


**(j)** enLL

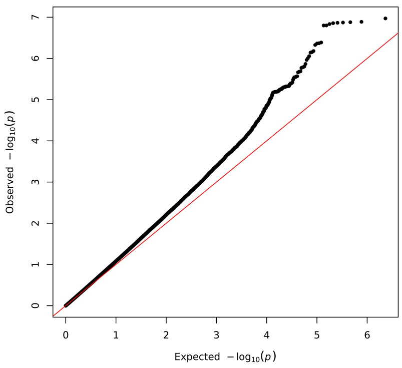


**(k)** enth

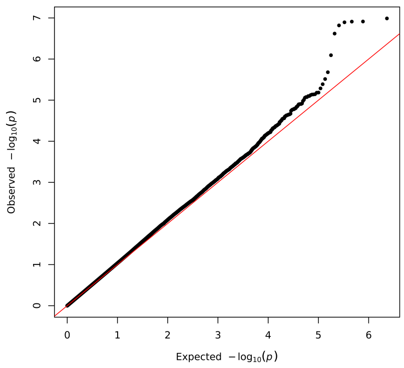


**(l)** FM

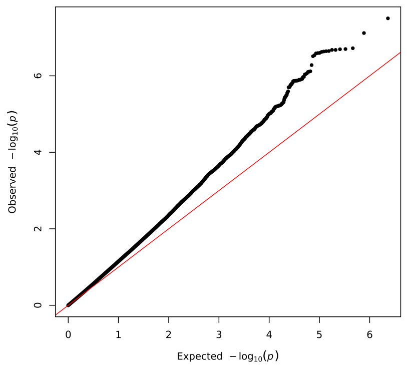


**(m)** gast

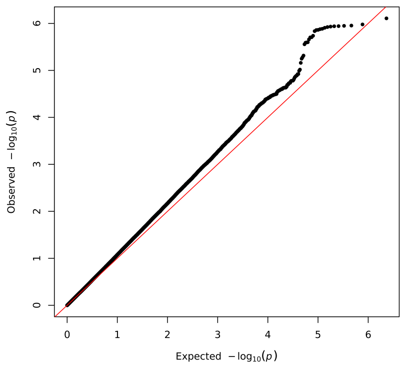


**(n)** CWP

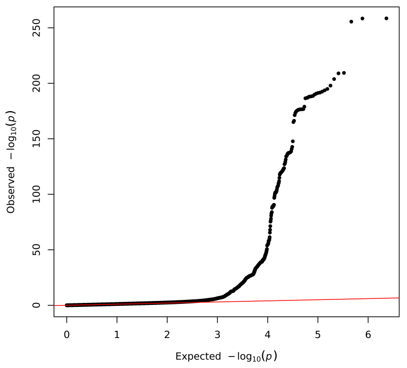


**(o)** gout

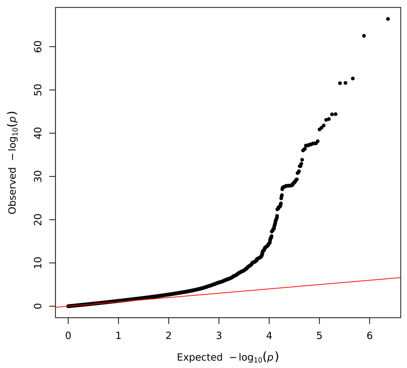


**(p)** hdch

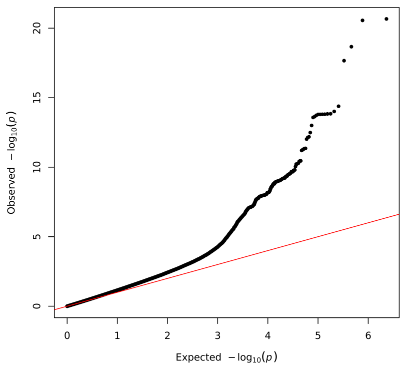


**(q)** hipA

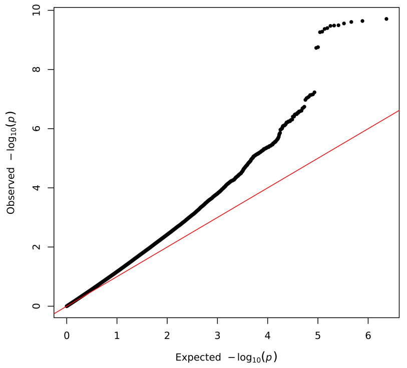


**(r)** hipP

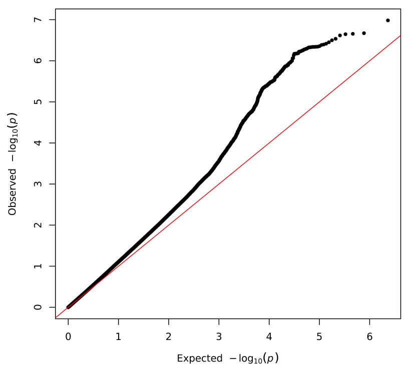


**(s)** IBS

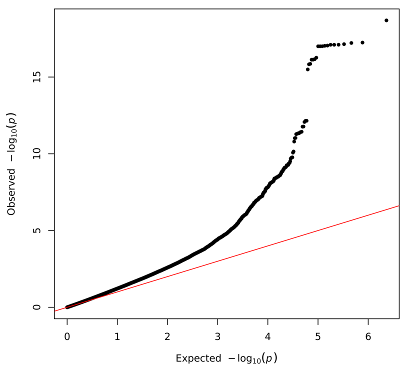


**(t)** kneA

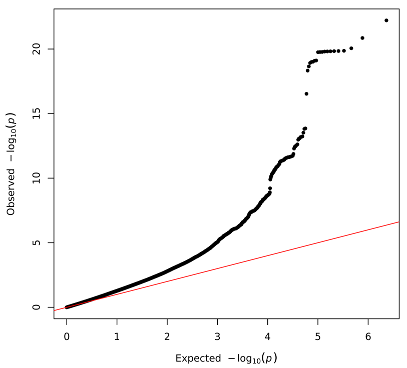


**(u)** kneP

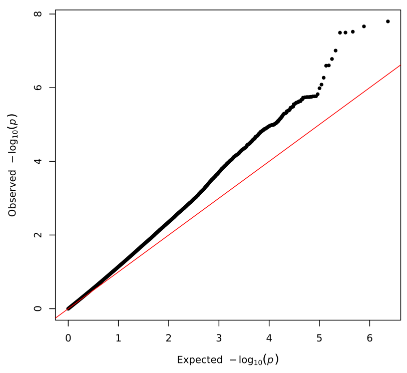


**(v)** legP

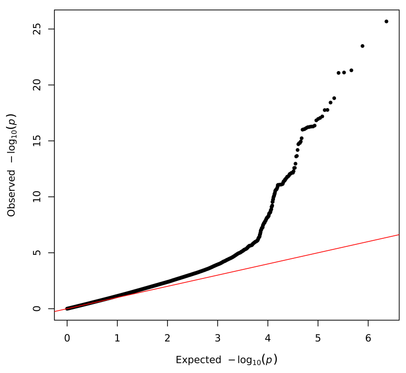


**(w)** mgrn

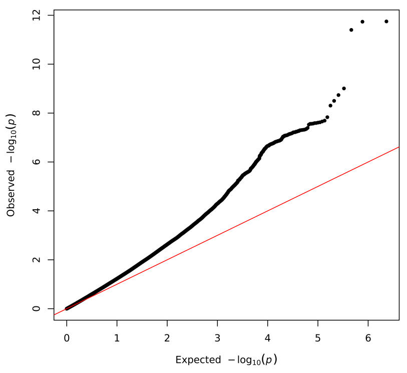


**(x)** neck

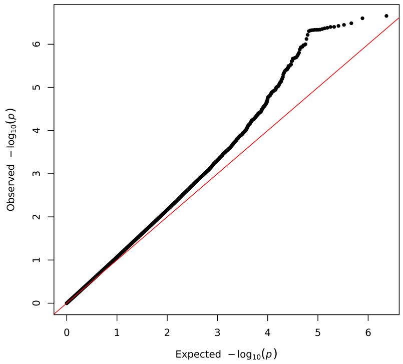


**(y)** oesp

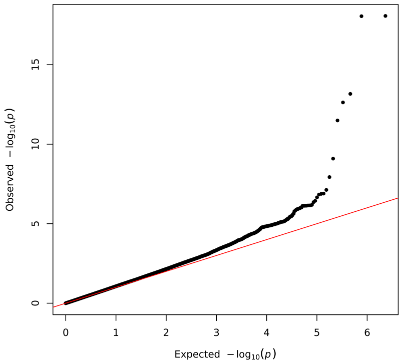


**(z)** rhAt

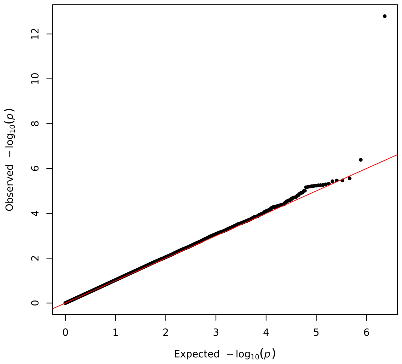


**(aa)** plrh

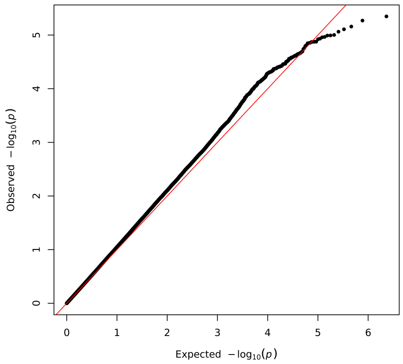


**(ab)** pnjt

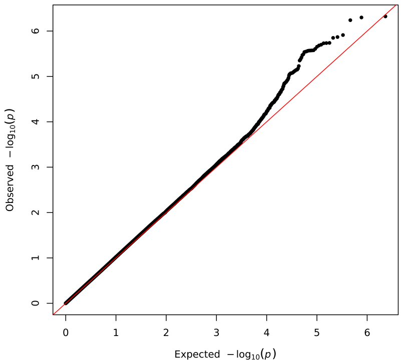


**(ac)** prst

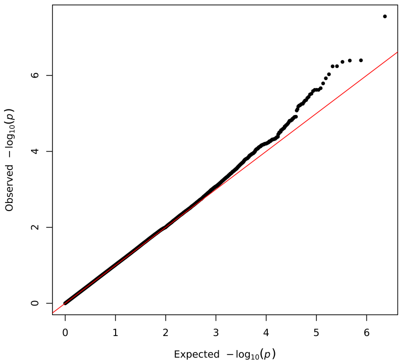


**(ad)** seRA

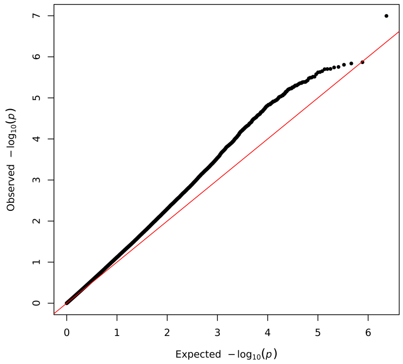


**(ae)** stmP

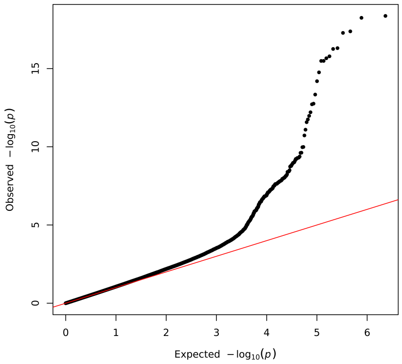


**(af)** ulcC

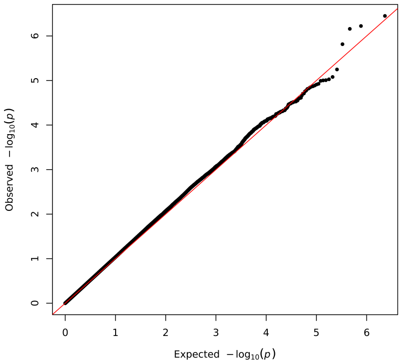


**(ag)** urCl

**Figure S2** Quantile-quantile (QQ) plots for 33 pain conditions. Condition definitions are in Table 1, and details are in Supplementary Table 1 ).

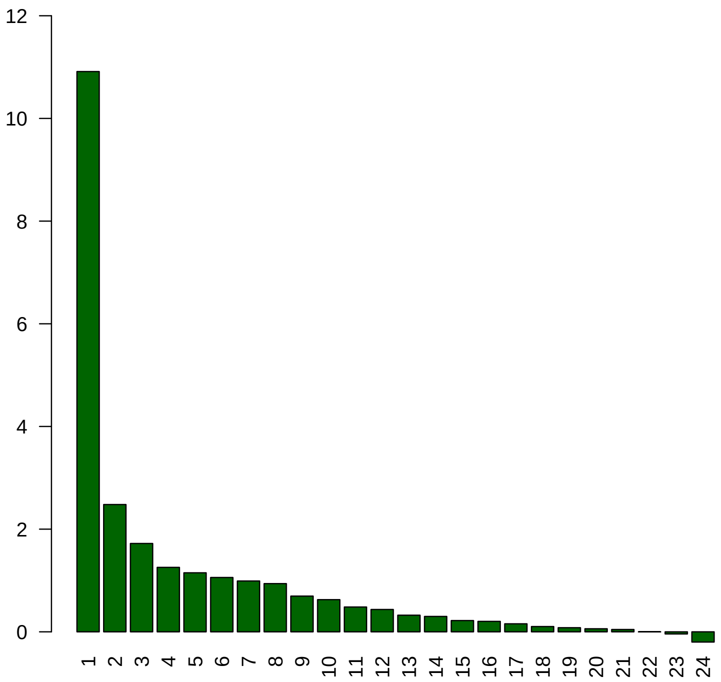


**Figure S3** Scree plot for 24 pain conditions (all autosomes), obtained using exploratory factor analysis on genetic correlations.

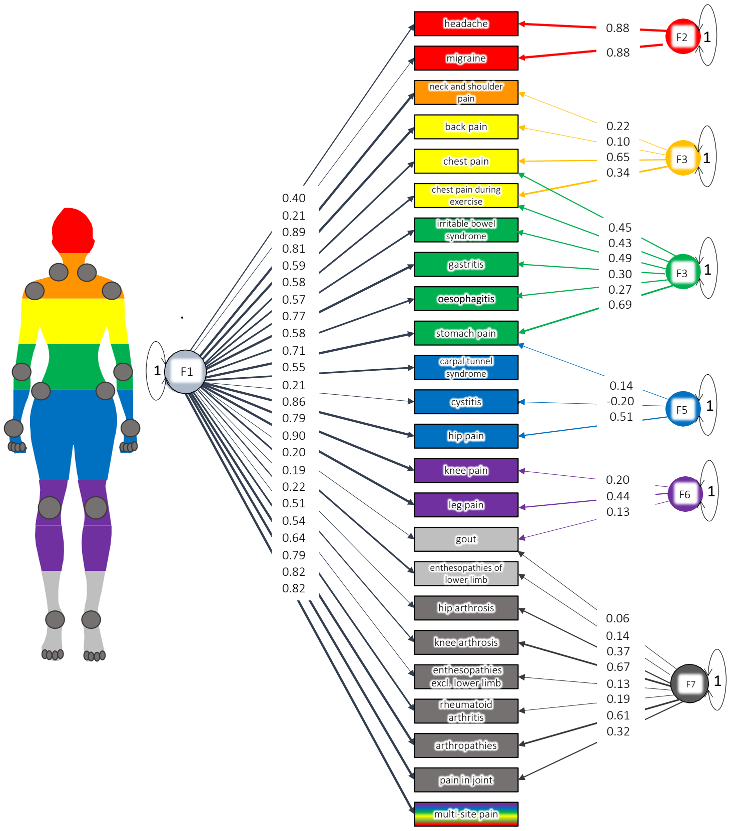


| | |
|---|---|
| CFI | 0.87 |
| SRMR | 0.087 |

**Figure S4** Structural equation model with hypothesis-driven anatomic groupings for 24 pain conditions. Factors: General (F1), Cranial (F2), Gastrointestinal (F3), Torso (F4), Pelvic (F5), Leg/Foot (F6) and Joint (F7). CFI, comparative fit index; SRMR, standardized root mean squared residual. All shown loadings are significant at $\alpha = 0.05$ except for all the conditions defining F6 "Leg/Foot": gout, knee pain, and leg pain; all the conditions defining F5 "Pelvic": cystitis, hip pain, and stomach pain; as well as enthesopathies of the lower limb and gout with F7 "Joint". (More information on all conditions in Supplementary Table 1).

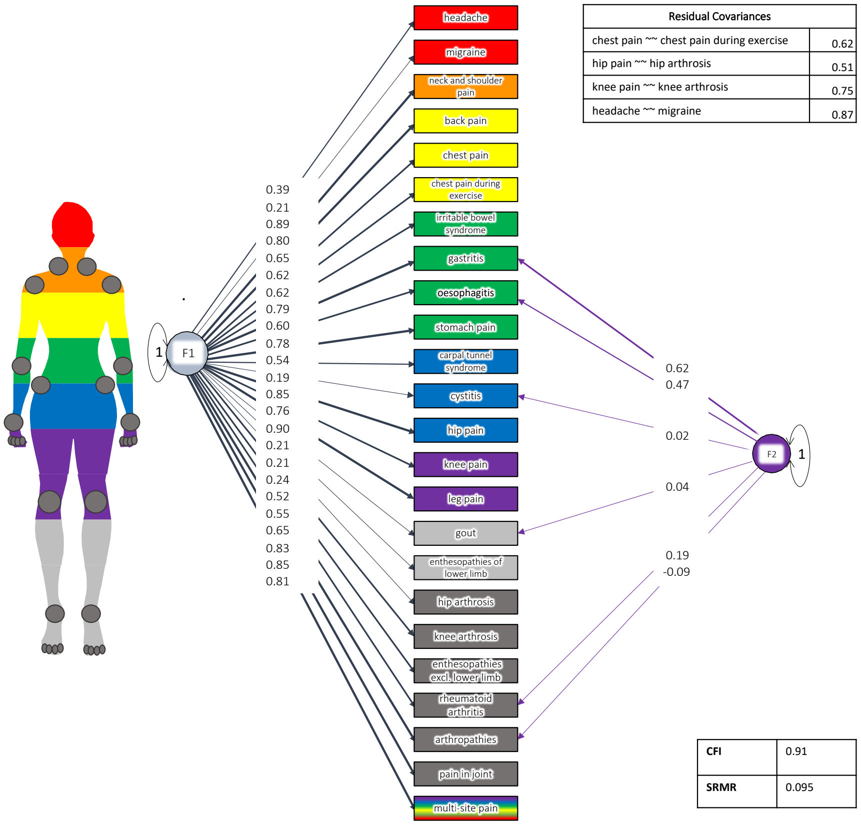


**Figure S5** Structural equation model with hypothesis-driven etiologic groupings for 24 pain conditions. Factors: General (F1), Inflammatory (F2). CFI, comparative fit index; SRMR, standardized root mean squared residual. All loadings are significant at $\alpha = 0.05$, except arthropathies, rheumatoid arthritis, cystitis, and gout on the "Inflammatory" factor. (More information on all conditions in Supplementary Table 1).

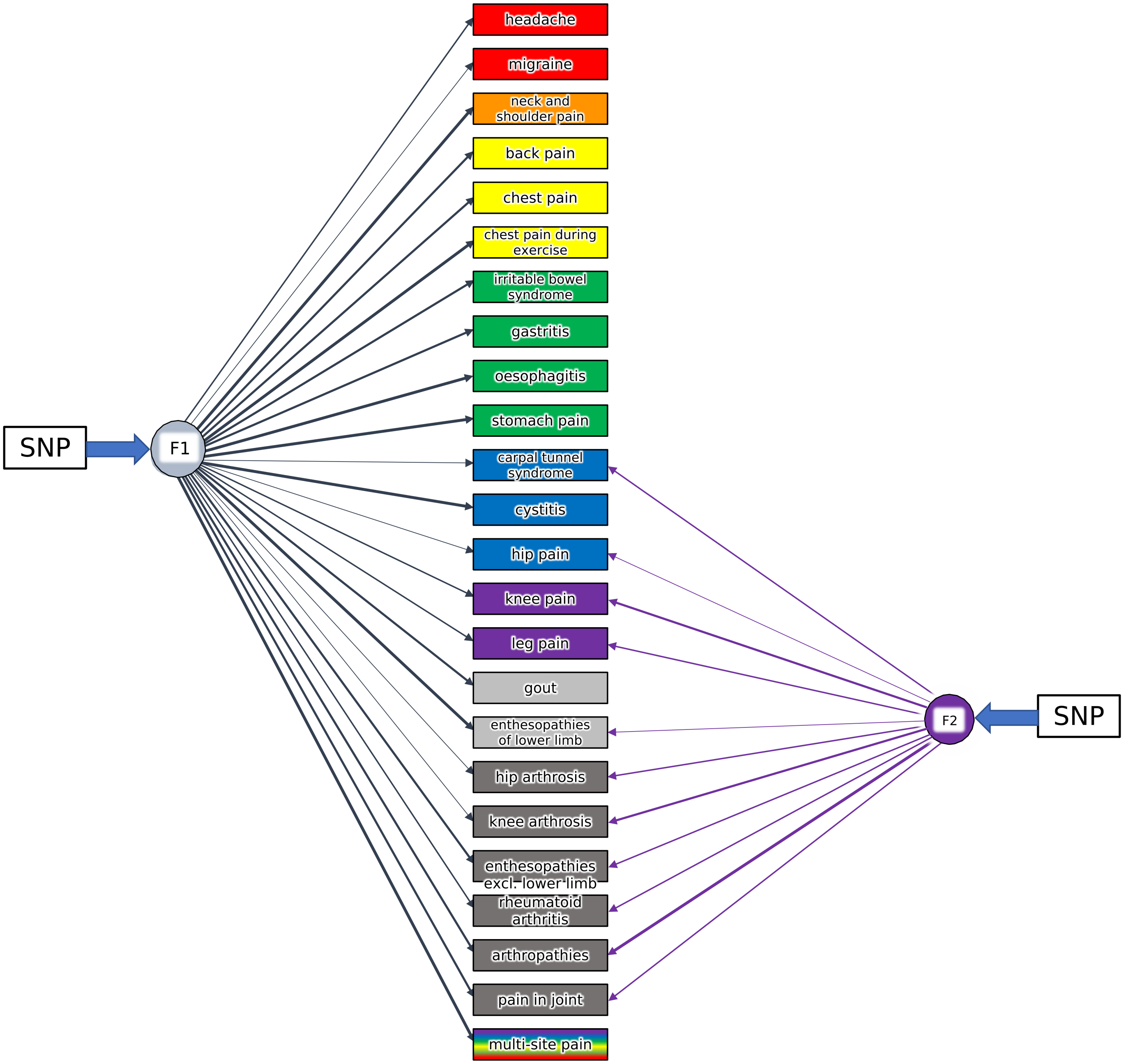


**Figure S6** The EFA-CFA model for 24 pain conditions, Figure 2A, with SNP effects. The pathway coefficients for each SNP were estimated for both factors in factor genome-wide association study (GWAS) in GenomicSEM. (More information on all conditions in Supplementary Table 1)

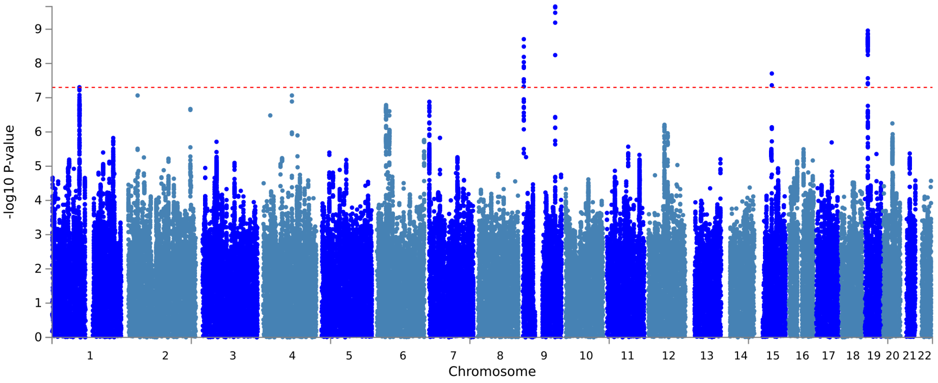


**(a)** Manhattan plot

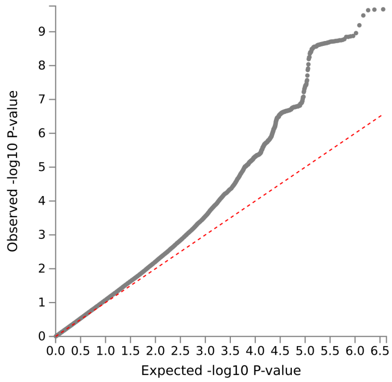


**(b)** QQ plot

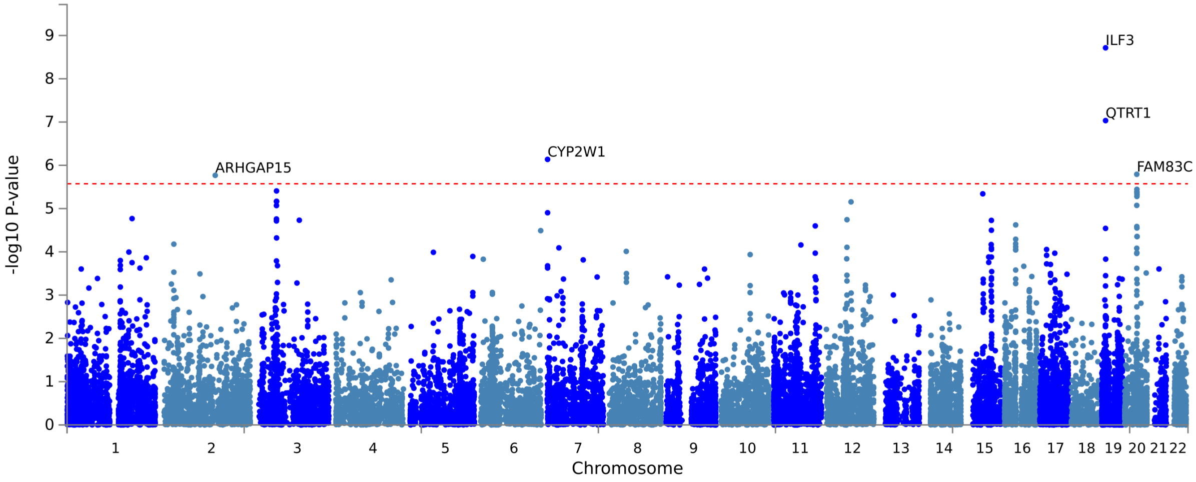


**(c)** Gene Manhattan plot

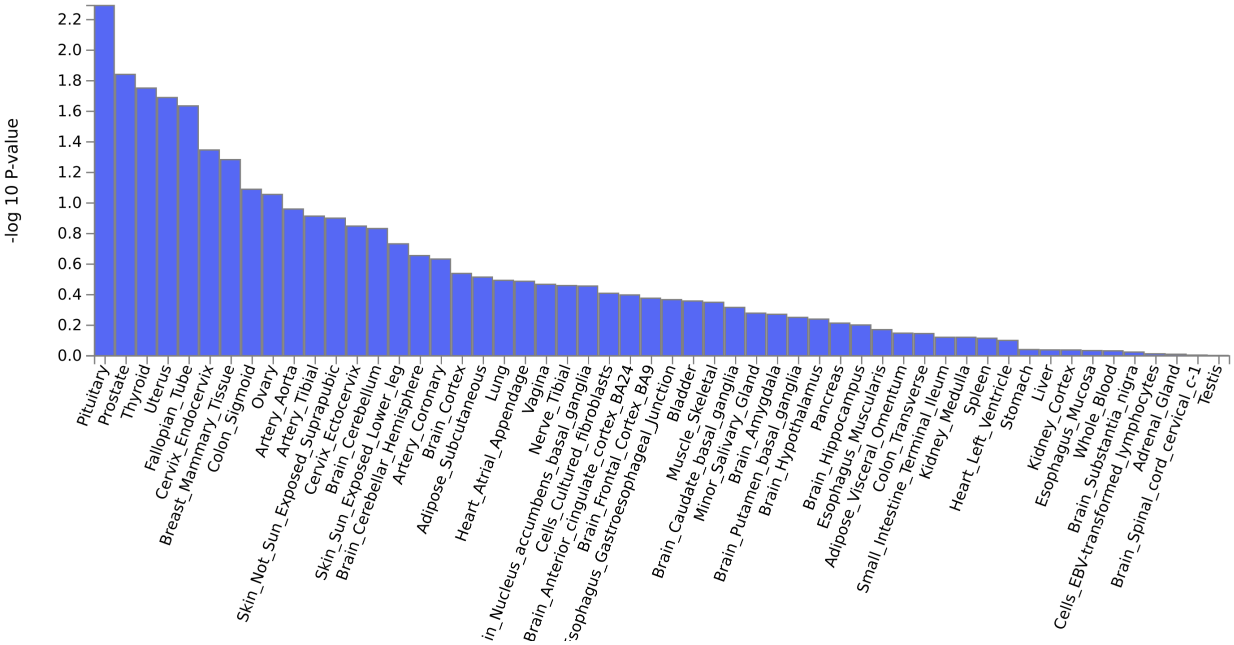


**(d)** 53 specific tissues

**Figure S7** Genome-wide association study (GWAS) results for the musculoskeletal pain factor (F2). SNP Manhattan (a) and quantile-quantile, QQ, (b) plots for F1 GWAS. (c) Gene-based genome-wide association Manhattan plot, with the top 31 associated genes labelled. (d) Gene property analysis for association between factor GWAS gene effects and gene expression levels in 53 specific tissues from GTEx, version 8.

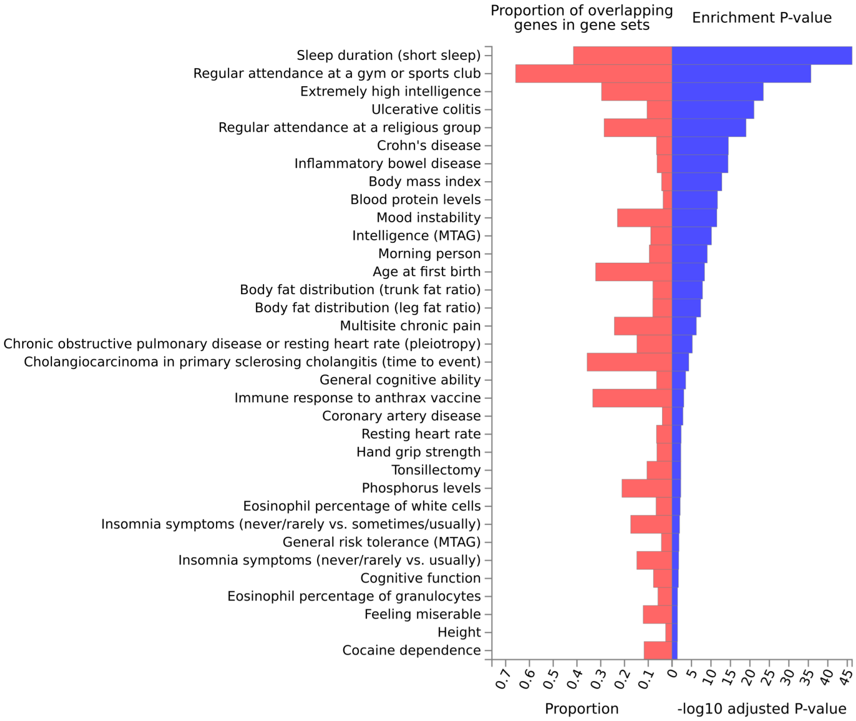


**Figure S8** FUMA plot of overlap of F1 genes with genes reported in previously published genome-wide association studies (GWAS).

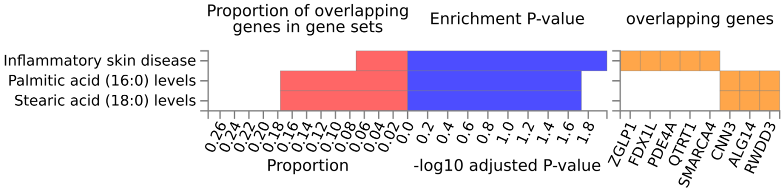


**Figure S9** FUMA plot of overlap of F2 genes with genes reported in previously published genome-wide association studies (GWAS).
